# *PKHD1L1* affects fertility in women

**DOI:** 10.64898/2026.08.20.26360790

**Authors:** Emmi Kapiainen, Minna K. Karjalainen, Petar B. Petrov, Riikka K. Arffman, Ulla Saarela, Sydney E. Parks, FinnGen, Eirini Trichia, Diego Aguilar-Ramirez, Lena Luyckx, Milena Myllykangas, Jason M. Torres, Jaime Berumen, Jesús Alegre-Díaz, Pablo Kuri-Morales, Roberto Tapia Conyer, Laura C. Cuello, Ramya P. Masand, Katri Pylkäs, Lari Lehtiö, Diana Monsivais, Terhi T. Piltonen, Johannes Kettunen, Renata Prunskaite-Hyyryläinen

## Abstract

Reproduction is one of the most fundamental biological processes in the human body, yet the molecules governing it remain incompletely understood. Here, we have characterized the role of *PKHD1L1* and its globally relatively common splice donor variant rs17368310 in female fertility. We demonstrate estrogen-responsive expression of PKHD1L1 in the human endometrial and Fallopian tube epithelium, identify the change in the rs17368310 mRNA sequence in endometrial tissue, and assess the possible effects of the variant on the PKHD1L1 protein through structural modeling. We reveal that women homozygous for rs17368310 have a persistently lower child count compared to other genotypes not only among all women but also among women who have undergone medical treatments for infertility in the Finnish population. We further show that rs17368310 associates with female infertility-related traits also in the Mexican population. These findings elucidate the effects of rs17368310 on fertility in millions of reproductive-age women across different populations.

## Introduction

Seamless function of the reproductive organs is critical for fertility. This includes successful ovulation, fertilization supported by the Fallopian tube, and the ability of the endometrium to correctly sense, receive, and sustain the implanting blastocyst, enabling later placental development and progression to term pregnancy. The World Health Organization estimates that ∼15% of all couples globally suffer from involuntary infertility^1^. Approximately 30–40% of all infertility cases can be solely attributed to the female partner, and up to 30% remain unexplained after excluding obvious gynecological and systemic diseases and impeding environmental factors^2,3^. Notably, many unexplained cases are suspected to depend on yet unidentified genetic causes^4,5^. Currently, most clinically relevant monogenic causes of female infertility are related to ovarian dysfunction^4–6^, but the accumulating data on the key molecular roles of the endometrium and Fallopian tubes for pregnancy establishment^7,8^ give reason to assume that gene variants disturbing their function are underrepresented in current data.

Genome-wide association studies (GWASs) utilizing data from large biobanks and population cohorts have recently identified associations at several loci for female infertility^9–11^. However, the biological mechanisms underlying these associations and the specific functions of the associated genetic variants are still largely unexplored. When following up the GWAS loci, identification of the causal variant responsible for the association signal can often be challenging, but in cases where the association signal includes a protein structure–changing variant, it has a high potential of being the variant causing the association. Recently, a strong recessive association of polycystic kidney and hepatic disease 1 like 1 (*PKHD1L1*) gene variant NC_000008.11:g.109459837G>C (rs17368310; NM_177531.6:c.7246+1G>C; NP_803875.2:p.(?)) with female infertility was identified^10^. As this variant affects a splice donor site, there is a very high likelihood for altered mRNA splicing and, therefore, a potential deleterious effect on the protein function (SpliceAI^12^ donor loss score 1.00, CADD score 35.0 [cadd.gs.washington.edu/snv, GRCh38-v1.7^13^]); however, the true molecular implications of this variant remain unknown.

PKHD1L1, also known as Fibrocystin-L, is a multi-domain, membrane-bound protein mostly residing in the extracellular space^14–16^. While its mechanism of action is yet unexplained, the involvement of PKHD1L1 in sustaining hearing ability as a component of the hair stereocilia machinery has, in recent years, been extensively demonstrated both directly in humans and by using mouse and zebrafish models^15–18^. Molecular level data on PKHD1L1 and female fertility is, at best, scarce—a handful of transcriptomic studies briefly mention potential changes of endometrial *PKHD1L1* expression in response to the menstrual cycle phase^19–22^, hormonal treatments^23,24^, endometriosis^25^ or recurrent implantation failure^26^, and suggest *PKHD1L1* expression in the bovine oviduct/human Fallopian tube^27,28^. In light of the abovementioned scattered reports, characterization of PKHD1L1 in female reproductive tract regulation urgently calls for more thorough studies.

Here, we set out to explore the molecular role of *PKHD1L1* and its splice donor variant rs17368310 in female fertility. We show that *PKHD1L1* is expressed in the human uterine and Fallopian tube epithelium and that rs17368310 disrupts *PKHD1L1* mRNA splicing with likely consequences on protein function. We further demonstrate that the variant has trans-ethnic associations with fertility-related traits that extend the impact of these findings to millions of women, in particular to those with Latin American ancestry. Collectively, our data show that *PKHD1L1* contributes significantly to fertility in women.

## Methods

### Human tissue material collection

Endometrial biopsies for RNA and venous blood samples for gDNA extraction were collected from women aged 21–42 years at the Oulu University Hospital, Oulu, Finland, as a part of a larger endPCOS study described in Lee *et al*. 2024^29^. Only healthy controls without PCOS were included in the present study. None of the study participants had been using hormonal medications for at least three months before study participation, they had no diseases or took no drugs that would affect the menstrual cycle, they had not been pregnant or breastfeeding within the last three months, and they were nonsmokers. The study was approved by the regional medical research ethics committee of the Northern Ostrobothnia Hospital District in Finland (EETTMK: 22/2013). All women signed informed consent. Venous blood samples (*n* = 63 women) were collected during study participation and stored in -20°C. The endometrial biopsies were collected using a plastic suction curette (Pipelle), snap-frozen in liquid nitrogen, and stored in -80°C. Proliferative phase endometrial samples (*n* = 4) were collected on menstrual cycle days 6–8 and secretory samples were timed with a luteinizing hormone (LH) surge measured from the urine (Clearblue Digital; Swiss Precision Diagnostics GmbH): early secretory samples, LH + 2–3 days (*n* = 6), mid-secretory samples, LH + 7–8 days (*n* = 8), and late secretory samples, LH + 11–12 days (*n* = 3). The presence of the corpus luteum was confirmed by ultrasound. Some women had donated samples in several menstrual cycle phases (different cycles), and some only one or no sample.

Endometrial (*n* = 9) and Fallopian tube (*n* = 2) tissues for immunohistochemistry were collected in 10% neutral buffered formalin and embedded in paraffin (FFPE tissue). Endometrial tissues (donor age 33–50 years) were obtained by a board-certified gynecological pathologist from an archived tissue bank at Baylor College of Medicine, Houston, TX, USA, under a protocol approved by the Institutional Review Board (IRB; H-21138). Menstrual cycle phases of the endometrial samples were assessed by a pathologist based on their histological appearance. Fallopian tubes were obtained following informed consent from donors (age 33–41 years) undergoing hysterectomy for pelvic pain (IRB H-21138). Histopathology analyses determined that the patients did not have endometriosis. Menstrual cycle phases of the Fallopian tube donors were not determined. All tissues were de-identified and obtained following the IRB-approved guidelines.

### Sequencing of genomic DNA

For classifying endometrial biopsy donors for RNA analyses, genomic DNA was extracted from venous blood samples using the DNeasy Blood & Tissue Kit (69504, Qiagen). The genomic region around the *PKHD1L1* rs17368310 variant was amplified by PCR using Phusion high-fidelity DNA polymerase (F530L, Thermo Fisher Scientific). Primer sequences are presented in **Table S1**. The thermocycling program consisted of an initial denaturation for 1 min at 98°C, 30 cycles of 98°C for 10 s/62°C for 30 s/72°C for 15 s, and a final extension at 72°C for 10 min. The PCR products were enzymatically purified using FastAP Thermosensitive Alkaline Phosphatase and Exonuclease I (EF0654 and EF0581, respectively; Thermo Fisher Scientific) according to the manufacturer’s instructions and sequenced using Sanger sequencing (ABI3500xL Genetic Analyzer system, Applied Biosystems) at Biocenter Oulu Sequencing Center, Finland, using the forward primers from the genotyping PCR. The sequencing data were processed using SnapGene software (version 8.0.1) or Chromas (version 2.6.6; Technelysium Pty Ltd).

For verifying the genotypes of FFPE tissue donors, slides were first deparaffinized and incubated in PBS at +4°C for 16 h. Genomic DNA was then isolated using the Quick-DNA Miniprep Plus Kit (D4068, Zymo Research) according to the manufacturer’s protocol for DNA isolation from FFPE samples. The genomic region around the *PKHD1L1* variant rs17368310 was amplified by PCR using Q5 High Fidelity DNA Polymerase (M0491S, New England BioLabs) according to the manufacturer’s protocol. Primer sequences are presented in **Table S1**. The thermocycling program consisted of an initial denaturation for 30 s at 98°C, 35 cycles of 98°C for 10 s/64°C for 30 s/72°C for 15 s, and a final extension at 72°C for 2 min. The PCR products were purified using the DNA Clean & Concentrator-5 Kit (D4003, Zymo Research) and sequenced by Genewiz in Houston, TX, USA, via Sanger sequencing using the forward primers from the genotyping PCR.

### RNA isolation and cDNA synthesis

RNA was extracted from the endometrial biopsies using the miRNeasy Mini kit (217004, Qiagen) and treated with RNase-free DNase I (EN0521, Thermo Fisher Scientific) according to the manufacturer’s protocol to remove any remains of genomic DNA. 1 µg of RNA was reverse transcribed using RevertAid First Strand cDNA synthesis kit (K1622, Thermo Fisher Scientific) and random hexamer oligos. cDNA was diluted 1:5 and used in RT-qPCR and in splicing analysis.

### RT-qPCR

Quantitative reverse transcription PCR (RT-qPCR) for endometrial *PKHD1L1* was performed using SsoAdvanced Universal SYBR® Green Supermix (1725274, Bio-Rad), a CFX96 Real-Time System (Bio-Rad), and *GAPDH* as a housekeeping gene. Primer sequences are presented in **Table S1**. The thermocycling program consisted of 40 cycles of 95°C for 30 s/60°C for 1 min, followed by a melt curve analysis. The donors of endometrial tissue were verified to not carry the rs17368310 variant allele (see chapter *Sequencing of genomic DNA*).

### Immunohistochemistry

After deparaffinization and rehydration, 5 µm thick FFPE tissue sections were boiled in 0.01 M sodium citrate (pH 6.0) for 20 min. Following antigen retrieval, sections were blocked with 3% H_2_O_2_ for 10 min at room temperature (RT), with avidin–biotin blocking kit (Vector Laboratories, SP-2001) according to the manufacturer’s protocol, and with 3% bovine serum albumin (BSA) in 1x Tris buffered saline with 0.1% Triton X-100 (TBST) for 1 h at RT. Sections were then incubated overnight at +4⁰C in mouse anti-PKHD1L1 antibody (immunogen NP_803875 aa 4105–4186, clone 1F5, catalogue number H00093035-M01, lot number K7241-1F5, Novus Biologicals) diluted 1:50 in 3% BSA in TBST. Negative control sections were incubated in the buffer without primary antibody. The PKHD1L1 antibody has previously been validated by showing positive staining in wild type and no staining in *Pkhd1l1* knockout mouse tissues^30^. Biotinylated goat anti-mouse IgG antibody (BA-9200, Vector Laboratories) diluted 1:200 in 3% BSA in TBST was then applied for 1 h at RT. Staining was visualized using first the VECTASTAIN Elite ABC-HRP kit (PK-6100, Vector Laboratories) as per the manufacturer’s instructions, followed by incubation in 125 µg/mL 3,3’-diaminobenzidine (DAB)/0.01% H_2_O_2_ in TBS for 8 min. Finally, the sections were counterstained with Harris hematoxylin, dehydrated, and mounted with Permount (Fisher Scientific). Stained sections were imaged with Leica Aperio AT2 DX System using a 40× objective, and the images were processed using Aperio ImageScope (Leica Biosystems). The donors of endometrial FFPE tissue were verified to not carry the rs17368310 variant allele (see chapter *Sequencing of genomic DNA*).

### Single-cell RNA sequencing analyses

scRNA-seq and cell type data were downloaded for endometrium and endometrial epithelium of non- pregnant uterus^31^ (https://www.reproductivecellatlas.org/), cancer-free and mostly premenopausal Fallopian tube^32^ (GEO: GSE151214), ampulla and fimbria of pre- and postmenopausal Fallopian tube^33^ (CELLxGENE: https://cellxgene.cziscience.com/collections/380ade76-e561-49a8-afb2-0f10b39c2c72), and adult ovary^34^ (GEO: GSE118127) to explore *PKHD1L1* expression. Analyses were done in R (v4.5, https://www.r-project.org/) with the tools from the Seurat package^35–38^. When needed, cells were filtered for 200< nGenes <2500 and mitochondrial genes <10%. When integration was required, RPCAIntegration was applied, following the analysis for Seurat v5 (https://satijalab.org/seurat/articles/seurat5_integration). All samples were integrated for the ovary data^34^ and the Fallopian tube data from Dinh *et al.* 2021^32^. For the Fallopian tube data from Weigert *et al.* 2025^33^, metadata from the study was used. For the endometrium, cell types were obtained from the contained metadata, whereas for the rest of the tissues, cell types were determined using Sc-type^39^ from the gene expression per cluster data provided by the respective study. Genes upregulated and downregulated (if provided) by at least 25% of cells per cluster were considered. Gene lists used were available as Supplementary Data 2 in Fan *et al.*, 2019^34^ and Tables S1 and S3 in Dinh *et al*., 2021^32^, for which epithelial cells were outlined as those expressing either *EPCAM*, *KRT7*, *KRT8*, *KRT18*, *OVGP1*, or *FOXJ1* as in the original study.

### Endometrial epithelial organoid RNA sequencing analysis

To study *PKHD1L1* expression in endometrial epithelial organoids, a previously reported RNA-seq dataset was utilized^40^. Briefly, endometrial epithelial cells were isolated from endometrial biopsies derived from regularly cycling, 27–39-year-old, overweight women, and cultured as organoids. Only controls without PCOS were included in the present analysis. Organoids were cultured without hormones (baseline), or 6 days in 10 nM estrogen (estrogen-treated), or 2 days in 10 nM estrogen and then 4 days in 10 nM estrogen, 1 µM progesterone, 0.25 mM cAMP, and 10 µM Wnt/β-catenin signaling inhibitor XAV-939 (a combination inducing decidualization), followed by RNA sequencing.

### In silico gene expression analysis

The Mammalian Reproductive Genetics Database V2^41^, containing organ-specific RNA-seq and digital PCR expression data, was accessed at https://orit.research.bcm.edu/MRGDv2, and a query for human *PKHD1L1* (Ensembl code ENSG00000205038, NCBI Gene ID 93035) in female tissues was performed.

### In silico alternative splicing analysis

The potential alternative splicing of *PKHD1L1* rs17368310 mRNA was explored using the openly available splicing prediction tools SpliceRover^42^ (http://bioit2.irc.ugent.be/rover/splicerover/), SpliceAI^12^ (https://spliceailookup.broadinstitute.org/), Pangolin^43^, and SpliceVault^44^ (https://kidsneuro.shinyapps.io/splicevault/). In SpliceRover, the query was performed using a sequence spanning from *PKHD1L1* exon 47 to exon 48 (containing the variant site) and the ‘human donors’ model. In SpliceVault, the query was performed for *PKHD1L1* donor 47, including all possible ranked events, cryptic positions, exon skipping events, and tissues.

### mRNA splicing analysis

For analyzing impaired splicing, endometrial cDNA samples from reference allele carriers (*n* = 3) and heterozygous rs17368310 variant carriers (*n* = 5) were amplified by PCR using Phusion high- fidelity DNA polymerase (F530L, Thermo Fisher Scientific) and primers located in exons 46 and 49 (**Table S1**). The thermocycling program consisted of an initial denaturation for 1 min at 98°C, 30 cycles of 98°C for 10 s/65°C for 20 s/72°C for 15 s, and a final extension at 72°C for 10 min. The products were separated on 4% agarose gel and compared to a molecular weight marker (SM0371, Thermo Fisher Scientific), and, under UV light, a 10 µL pipette tip was used to gently collect DNA only from a specific band onto the tip. DNA from the tip was eluted to 10 µL water by heating at 80°C for 5 min, and a second PCR with the same primer set was carried out to amplify only the desired band. The success of the second PCR was verified on agarose gel, and enzymatic purification and Sanger sequencing was performed as for the genomic DNA described above (see chapter *Sequencing of genomic DNA*). For analyzing potential exon skipping events impacting a wider region, primers located in exons 45 and 51 were used in PCR (5% DMSO in the final reaction), and the products were separated on 3% agarose gel and compared to a molecular weight marker (SM0311, Thermo Fisher Scientific). This thermocycling program consisted of an initial denaturation for 1 min at 98°C, 35 cycles of 98°C for 10 s/63°C for 30 s/72°C for 50 s, and a final extension at 72°C for 10 min. Endometrial cDNA samples from both control and heterozygous women were used for sequencing regardless of their menstrual cycle phase but were matched with a corresponding phase reference in the gel runs. The HGVS Nomenclature–aligning descriptions were checked using Mutalyzer, version 3.2.0.dev0^45^ (https://mutalyzer.nl/).

### In silico protein analyses

Human PKHD1L1 protein domain composition was obtained by combining the search results of UniProt (Q86WI1), NCBI (NP_803875.2), and SMART (http://smart.embl-heidelberg.de/). Potential for general proprotein convertase cleavage was analyzed with the ProP 1.0 tool^46^ (https://services.healthtech.dtu.dk/services/ProP-1.0/). Previous publications reporting the composition or proteolytic processing of the human PKHD1L1 protein were also used as a reference^14,16,17,47^.

PKHD1L1 protein multiple sequence alignment was performed with MUSCLE3^48^ available at EMBL-EBI (https://www.ebi.ac.uk/jdispatcher/msa/muscle?stype=protein) for selected mammals. The NCBI accession numbers of the sequences are listed in **Table S2**. Signal sequences were included in the analysis, and the human amino acid sequence was used as a residue numbering reference. The alignment was visualized in the Jalview program^49^ using the “Color by Annotation” and “Conservation” coloring function.

Structural modeling of the first TMEM2-like region of PKHD1L1 was carried out using AlphaFold Server (AlphaFold 3)^50,51^ using the residues Phe2184–Ala2840 (NCBI reference sequence NP_803875.2 on December 4^th^, 2024). To generate a conserved molecular surface for the fragment, the full human sequence was used to identify similar proteins by a Blastp search from a ClusteredNR database^52^. After ensuring multiple entries of isoforms from the same species were excluded, a multiple sequence alignment of the resulting 87 entries was generated with Cobalt^53^. The NCBI accession numbers of the sequences are listed in **Table S2**. The multiple sequence alignment was annotated for conservation using Consurf server^54^ and the conservation scores were mapped to the predicted model of human PKHD1L1. The crystal structure of TMEM2 was obtained from PDB (ID 8C6I)^55^. Visualization of the structures was done with PyMOL (The PyMOL Molecular Graphics System, Version 2.5.0, Schrödinger, LLC).

Potential PTMs on the amino acid insertion ANFSSSR (novel residues inserted between Thr2415– Gly2416) and the immediate flanking sequence of human PKHD1L1 protein as a result of variant rs17368310 were explored using The Eukaryotic Linear Motif (ELM) resource^56^ (http://elm.eu.org/), MusiteDeep^57^ (https://www.musite.net/), NetNGlyc 1.0^58^ (https://services.healthtech.dtu.dk/services/NetNGlyc-1.0/), NetOGlyc 4.0^59^ (https://services.healthtech.dtu.dk/services/NetOGlyc-4.0/), and NetPhos 3.1^60,61^ (https://services.healthtech.dtu.dk/services/NetPhos-3.1/).

### The FinnGen study cohort and data analyses

Data from the FinnGen study was used (1) to analyze the association of *PKHD1L1* locus genetic variants with medical treatment for female infertility, and (2) to assess the association of *PKHD1L1* variant rs17368310 with the number of children in women. The FinnGen study is a large-scale genomics initiative that has analyzed over 500,000 Finnish biobank samples and correlated genetic variation with national health register data to understand disease mechanisms and predispositions. The project is a collaboration between research organizations and biobanks within Finland and international industry partners^62^. Study subjects in FinnGen have provided informed consent for biobank research based on the Finnish Biobank Act. Alternatively, separate research cohorts established prior to the Finnish Biobank Act coming into effect (in September 2013) and the start of FinnGen (August 2017) were collected based on study-specific consents and later transferred to the Finnish biobanks after approval by Fimea (Finnish Medicines Agency), the National Supervisory Authority for Welfare and Health. Recruitment protocols followed the biobank protocols approved by Fimea. The Coordinating Ethics Committee of the Hospital District of Helsinki and Uusimaa (HUS) statement number for the FinnGen study is Nr HUS/990/2017.

The FinnGen study is approved by Finnish Institute for Health and Welfare (THL/2031/6.02.00/2017, THL/1101/5.05.00/2017, THL/341/6.02.00/2018, THL/2222/6.02.00/2018, THL/283/6.02.00/2019, THL/1721/5.05.00/2019 and THL/1524/5.05.00/2020), Digital and population data service agency (VRK43431/2017-3, VRK/6909/2018-3, VRK/4415/2019-3), the Social Insurance Institution (KELA 58/522/2017, KELA 131/522/2018, KELA 70/522/2019, KELA 98/522/2019, KELA 134/522/2019, KELA 138/522/2019, KELA 2/522/2020, KELA 16/522/2020), Findata (THL/2364/14.02/2020, THL/4055/14.06.00/2020, THL/3433/14.06.00/2020, THL/4432/14.06/2020, THL/5189/14.06/2020, THL/5894/14.06.00/2020, THL/6619/14.06.00/2020, THL/209/14.06.00/2021, THL/688/14.06.00/2021, THL/1284/14.06.00/2021, THL/1965/14.06.00/2021, THL/5546/14.02.00/2020, THL/2658/14.06.00/2021, THL/4235/14.06.00/2021), Statistics Finland (TK-53-1041-17 and TK/143/07.03.00/2020 (earlier TK-53-90-20), TK/1735/07.03.00/2021, TK/3112/07.03.00/2021) and Finnish Registry for Kidney Diseases (permission/extract from the meeting minutes on 4th July 2019).

The Biobank Access Decisions for FinnGen samples and data utilized in FinnGen Data Freeze 12 include: THL Biobank BB2017_55, BB2017_111, BB2018_19, BB_2018_34, BB_2018_67, BB2018_71, BB2019_7, BB2019_8, BB2019_26, BB2020_1, BB2021_65, Finnish Red Cross Blood Service Biobank 7.12.2017, Helsinki Biobank HUS/359/2017, HUS/248/2020, HUS/430/2021 §28, §29, HUS/150/2022 §12, §13, §14, §15, §16, §17, §18, §23, §58, §59, HUS/128/2023 §18, Auria Biobank AB17-5154 and amendment #1 (August 17 2020) and amendments BB_2021-0140, BB_2021-0156 (August 26 2021, Feb 2 2022), BB_2021-0169, BB_2021-0179, BB_2021-0161, AB20-5926 and amendment #1 (April 23 2020) and its modifications (Sep 22 2021), BB_2022-0262, BB_2022-0256, Biobank Borealis of Northern Finland_2017_1013, 2021_5010, 2021_5010 Amendment, 2021_5018, 2021_5018 Amendment, 2021_5015, 2021_5015 Amendment, 2021_5015 Amendment_2, 2021_5023, 2021_5023 Amendment, 2021_5023 Amendment_2, 2021_5017, 2021_5017 Amendment, 2022_6001, 2022_6001 Amendment, 2022_6006 Amendment, 2022_6006 Amendment, 2022_6006 Amendment_2, BB22-0067, 2022_0262, 2022_0262 Amendment, Biobank of Eastern Finland 1186/2018 and amendment 22§/2020, 53§/2021, 13§/2022, 14§/2022, 15§/2022, 27§/2022, 28§/2022, 29§/2022, 33§/2022, 35§/2022, 36§/2022, 37§/2022, 39§/2022, 7§/2023, 32§/2023, 33§/2023, 34§/2023, 35§/2023, 36§/2023, 37§/2023, 38§/2023, 39§/2023, 40§/2023, 41§/2023, Finnish Clinical Biobank Tampere MH0004 and amendments (21.02.2020 & 06.10.2020), BB2021-0140 8§/2021, 9§/2021, §9/2022, §10/2022, §12/2022, 13§/2022, §20/2022, §21/2022, §22/2022, §23/2022, 28§/2022, 29§/2022, 30§/2022, 31§/2022, 32§/2022, 38§/2022, 40§/2022, 42§/2022, 1§/2023, Central Finland Biobank 1-2017, BB_2021-0161, BB_2021-0169, BB_2021- 0179, BB_2021-0170, BB_2022-0256, BB_2022-0262, BB22-0067, Decision allowing to continue data processing until 31st Aug 2024 for projects: BB_2021-0179, BB22-0067,BB_2022-0262, BB_2021-0170, BB_2021-0164, BB_2021-0161, and BB_2021-0169, and Terveystalo Biobank STB 2018001 and amendment 25th Aug 2020, Finnish Hematological Registry and Clinical Biobank decision 18th June 2021, Arctic biobank P0844: ARC_2021_1001.

Genetic and clinical data from FinnGen Data Release R12 were used. Affymetrix ThermoFisher Axiom custom and Illumina GWAS arrays were used for SNP genotyping of the FinnGen samples.

Variants violating the Hardy-Weinberg equilibrium (HWE p-value < 1 × 10-6), and those with minor allele count <3 or high missingness (>2%) were excluded. Samples with high heterozygosity (±3 SD), non-Finnish ancestry, non-matching sex, or high genotype missingness (>2%) were excluded. Imputation of SNP genotypes was performed using Beagle 4.1; a population-specific SISu v4.2 reference panel was used. Variants with imputation info score <0.6 were excluded.

In the genetic association analysis, women with at least three purchases of medical treatments for infertility were defined as cases (*n* = 9,381) based on the Social Insurance Institution (KELA) registry of medicine purchases. Women with purchases of the following ATC codes were included: G03GA01, G03GA02, G03GA04, G03GA05, G03GA06, G03GA09, G03GA10, G03GA30, and G03GB02 (**Table S3**). Non-case women (*n* = 272,683) were used as controls. Association analysis was performed under the additive and recessive models using whole-genome regression with REGENIE v. 2.2.4^63^; age, 10 principal components, and genotyping batches were included as covariates. Significance was considered at the common threshold from genome-wide significance *p*<5 x 10^-8^. The Ensembl Variant Effect Predictor^64^ was used to infer functional consequences of genetic variants. In a phenome-wide association look-up, previous associations of rs17368310 were investigated from the GWAS Catalog^65^ and FinnGen R12 endpoints^62^.

In the analysis of the number of children, data extracted from the Finnish Population and Medical Birth Registries were utilized. To include only women who had reached the age of 45 years and for whom reliable information regarding the number of children was available through the registries, the analyzed women were restricted to those born between 1938 and 1978 (*n*=186,631). The differences in the mean number of children and in genotype counts between women with and without children were tested using an independent samples t-test and the χ2 test, respectively, with R, v. 4.3.1.

### The Mexico City Prospective Study (MCPS) study cohort and data analyses

Genetic data and questionnaire-based information from the MCPS were used to assess the association of *PKHD1L1* rs17368310 with the number of pregnancies. The MCPS is a cohort of ∼150,000 participants recruited at 35 years of age or older in Mexico City from 1998 to 2004, of whom 66% are female^66^. At recruitment, participants provided a blood sample and completed a standardized health questionnaire. Data from 94,113 women (rs17368310 genotypes: 44,929 G/G, 39,547 C/G, 9,637 C/C) representing mixed ancestries (genetically most representative of indigenous American origin) were available.

The MCPS was approved by the Mexican Ministry of Health, the Mexican National Council of Science and Technology (0595 P-M), the Central Oxford Research Ethics Committee (C99.260), and the Ethics and Research Commissions of the Faculty of Medicine at the National Autonomous University of Mexico (FMED/CI/SPLR/067/2015). Transport and long-term storage of blood samples (plasma and buffy coat) at the Clinical Trial Service Unit sample archive, University of Oxford, were approved by the Mexican Ministry of Health. All participants provided written informed consent.

As previously described for the MCPS cohort^67^, genotyping was performed with the Illumina Infinium Global Screening Array (GSA) v2.0 at the Regeneron Genetics Center (RGC) Sequencing Lab, generating a dataset of 650,381 variants for 140,831 participants. For this analysis, the QC workflow was modified by performing genotype missingness filtering prior to individual missingness filtering and applying an individual missingness threshold of 0.10. The resulting quality-controlled genotyping array dataset included 539,448 autosomal variants and 18,560 chromosome X variants for 140,829 individuals. Imputation was performed using the TOPMed Imputation Server and the TOPMed version r2 reference panel of 97,256 whole genome sequenced samples that includes 308,107,085 autosomal and chromosome X variants.

Genetic association testing under the additive and recessive models was conducted using REGENIE among 94,113 women with complete information on the number of pregnancies and genetic data after exclusion of unreliable linkage of genetic to phenotypic data and potential X/Y anomalies. The covariates included were age, age squared, district of residence, and the first seven genetic principal components. For the recessive model, REGENIE’s flag ‘--test recessive’ was used. The phenotype used was self-reported number of pregnancies among female MCPS participants (https://datashare.ndph.ox.ac.uk/mexico/field.cgi?id=59). The phenotype was normalized using the ‘--apply-rint’ flag in REGENIE. The difference in the mean number of pregnancies was additionally tested using an independent samples t-test with R, v. 4.3.1.

### Statistical analyses and data presentation

The FinnGen and MCPS data were analyzed as described above in their respective chapters. Shapiro– Wilk test was used to confirm normal distribution of the data (GraphPad Prism 10), Levene’s test to analyze the homogeneity of variances (OriginPro 2025, OriginLab), and Welch’s ANOVA followed by Dunnett’s T3 multiple comparisons test (Prism) to compare the means in endometrial expression analyses. Data are presented as individual values with mean±SD with the exact *p*-values shown when *p* ≤ 0.05, or as mean±95% confidence interval (CI), as indicated in the corresponding figure legends or tables. The exact number of biological samples used in each experiment or cohort participants is indicated in the corresponding figure legends or tables, respectively.

## Results

### PKHD1L1 is expressed in an estrogen-dependent manner in the female reproductive tract epithelium

Despite the proposed association of *PKHD1L1* with women’s infertility, its expression pattern in the female reproductive tissues has not been reported in sufficient detail. The endometrium of menstruating women is constantly remodeled in response to hormonal fluctuation and can be roughly divided into proliferative (follicular) and early, mid-, and late secretory (luteal) phases (**Fig. 1A**)^68^. Compellingly, the openly available Mammalian Reproductive Genetics Database V2 (MRGDv2)^41^ suggested considerably high *PKHD1L1* expression levels specifically in the early secretory endometrium (**Fig. S1A**). Using endometrial tissue biopsies and RT-qPCR we assessed that endometrial *PKHD1L1* expression levels indeed alter at different phases of the menstrual cycle, strikingly peaking at the early secretory phase and being the lowest in the late secretory phase (**Fig. 1B**). Our immunostaining of endometrial tissue localized PKHD1L1 protein explicitly into the epithelium, both luminal and glandular (**Fig. 1C and Fig. S1B**), and the staining intensity in different cycle phases followed similar trends as the RT-qPCR data (**Fig. 1B**). We then leveraged a published single-cell RNA-sequencing (scRNA-seq) dataset^31^ to gain even deeper understanding of the specific endometrial cell types expressing *PKHD1L1*. This confirmed highly epithelial-specific expression pattern in both the endometrium and endocervix, along with a small cluster of *PKHD1L1*+ lymphatic endothelium (**Fig. 1D**). *PKHD1L1* expression was by far the most prominent in the endometrial pre-luminal and pre-glandular epithelial cells (**Fig. 1D**), which are transitory cell states found only during the early secretory phase^31^. Proliferative phase epithelia displayed in general modest and mid- and late secretory phase epithelia low *PKHD1L1* expression levels (**Fig. 1D**). Nevertheless, *PKHD1L1* expression was not solely restricted to one epithelial cell subtype but was found in both luminal, glandular, ciliated, and *SOX9*+ progenitor epithelial cells^31^ (**Fig. 1D**). All the endometrial expression datasets align well with each other and with previous transcriptomic studies^19,20,22^, any slight discrepancies likely being a result of variation in the sample collection timing within the window of each menstrual cycle phase. The timing of endometrial PKHD1L1 expression peak suggested that it may be induced by rising estrogen levels (**Fig. 1A–D**). The analysis of RNA-seq data of cultured human endometrial epithelial organoids^40^ revealed a strong upregulation of *PKHD1L1* transcription upon treatment with solely estrogen compared to non-treated cells (baseline), while a cocktail of estrogen, progesterone, cAMP, and Wnt/β-catenin signaling inhibitor, which mimics the secretory phase and induces decidualization, did not generate as significant upregulating effect (**Fig. 1E**).

**Figure 1.**
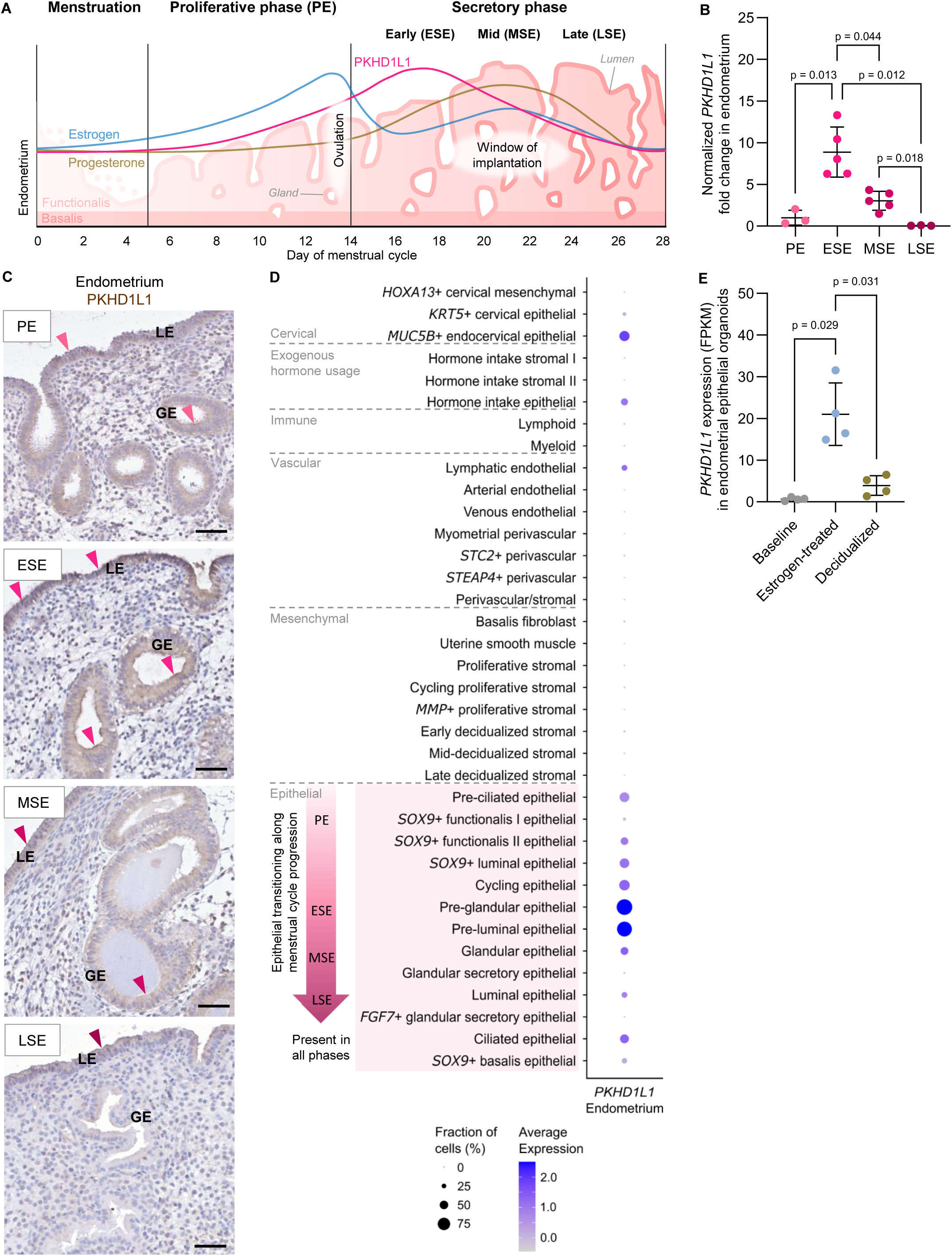
PKHD1L1 is expressed in the human endometrial epithelium. **A**, A graphical presentation of the endometrial remodeling across the human menstrual cycle. The pink line summarizes the course of PKHD1L1 expression based on the data presented in panels **B–E**. **B**, RT-qPCR analysis of *PKHD1L1* expression in endometrial samples from different menstrual cycle phases. *n* = 3 PE and LSE and *n* = 5 ESE and MSE samples. Within one cycle phase, all samples are from different women; across the cycle phases, some samples are derived from the same women. **C**, PKHD1L1 immunostaining of the endometrium in different menstrual cycle phases. Arrowheads mark the most intense signal. Scale bar 50 µm; *n* = 2 PE, *n* = 3 ESE, *n* = 3 MSE, and *n* = 1 LSE samples. **D**, scRNA-seq analysis of *PKHD1L1* expression in uterine cells, analyzed using the dataset of ***Marečková et al. 2024***. **E**, RNA-seq analysis of *PKHD1L1* expression in human endometrial epithelial organoids at baseline and when stimulated with estrogen alone or with decidualization- inducing compounds (estrogen, progesterone, cAMP, and Wnt/β-catenin signaling inhibitor (***Luyckx et al. 2025***; *n* = 4). Welch’s ANOVA followed by Dunnett’s T3 post hoc test was applied in **B** and **E**; data is presented as individual values with mean±SD. ESE, early secretory endometrium; FGF7, fibroblast growth factor 7; GE, glandular epithelium; HOXA13, homeobox A13; KRT, keratin; MMP, matrix metalloprotease; MSE, mid-secretory endometrium; MUC5B, mucin 5B; LE, luminal epithelium; LSE, late secretory endometrium; PE, proliferative endometrium; SOX9, SRY-box transcription factor 9; STC2, stanniocalcin 2; STEAP4, STEAP4 metalloreductase.

The Fallopian tubes act as an important bridge between the ovaries and the uterus, fine-tuning the molecular environment to favor fertilization (**Fig. 2A**). The data in the MRGDv2 database hinted towards significant *PKHD1L1* expression in the Fallopian tubes (**Fig. S1A**). Considering this, we performed immunohistochemistry and detected PKHD1L1 protein in the Fallopian tube epithelium (**Fig. 2B and Fig. S1C**). To complement our available samples, we made use of accessible scRNA- seq data^32^. Distinct *PKHD1L1* expression was observed in a subset of oviductal glycoprotein 1 (OVGP1)–positive secretory epithelial cells (**Fig. 2C**). These *OVGP1*+ secretory cells (secretory cell cluster 2^32^) have been suggested to be secretory cell progenitors^69,70^ and are directly hormonally regulated as evident from their high levels of both progesterone and estrogen receptors and the drastic decrease of this cell type after menopause^33,70^. *OVGP1* is specifically expressed during the late proliferative and very early secretory phase in the Fallopian tube epithelium^33,71–73^, indicating that *PKHD1L1* expression is also timed around ovulation. This was further supported by the observation that *PKHD1L1* expression is predominantly responsive to increasing estrogen level (**Fig. 1E**), which directly precedes ovulation (**Fig. 1A**), as well as another Fallopian tube scRNA-seq dataset^33^ showing that *PKHD1L1* levels plummet during the secretory phase (all secretory phases pooled) and at post- menopause (**Fig. S1D**). Within the Fallopian tube, *PKHD1L1* expression was observed both in the oocyte-capturing fimbriae, the bridging infundibulum, and the ampulla, the optimal site of fertilization (**Fig. 2A and B, Fig. S1D**). Finally, conversely to the endometrium and Fallopian tubes and in line with the MRGDv2 data (**Fig. 1B–D**, **Fig. 2B and C, Fig. S1A**), scRNA-seq data^34^ showed very low ovarian *PKHD1L1* expression, restricted only to a small subset of lymphatic endothelial cells (**Fig. 2D**). Moreover, the early pregnancy decidua and placenta were negative for expression according to the MRGDv2 data (**Fig. S1A**). Collectively, these results establish PKHD1L1 as a marker of the endometrial and Fallopian tube epithelium during the potential fertilization time window (**Figs. 1 and 2, Fig. S1**).

**Figure 2.**
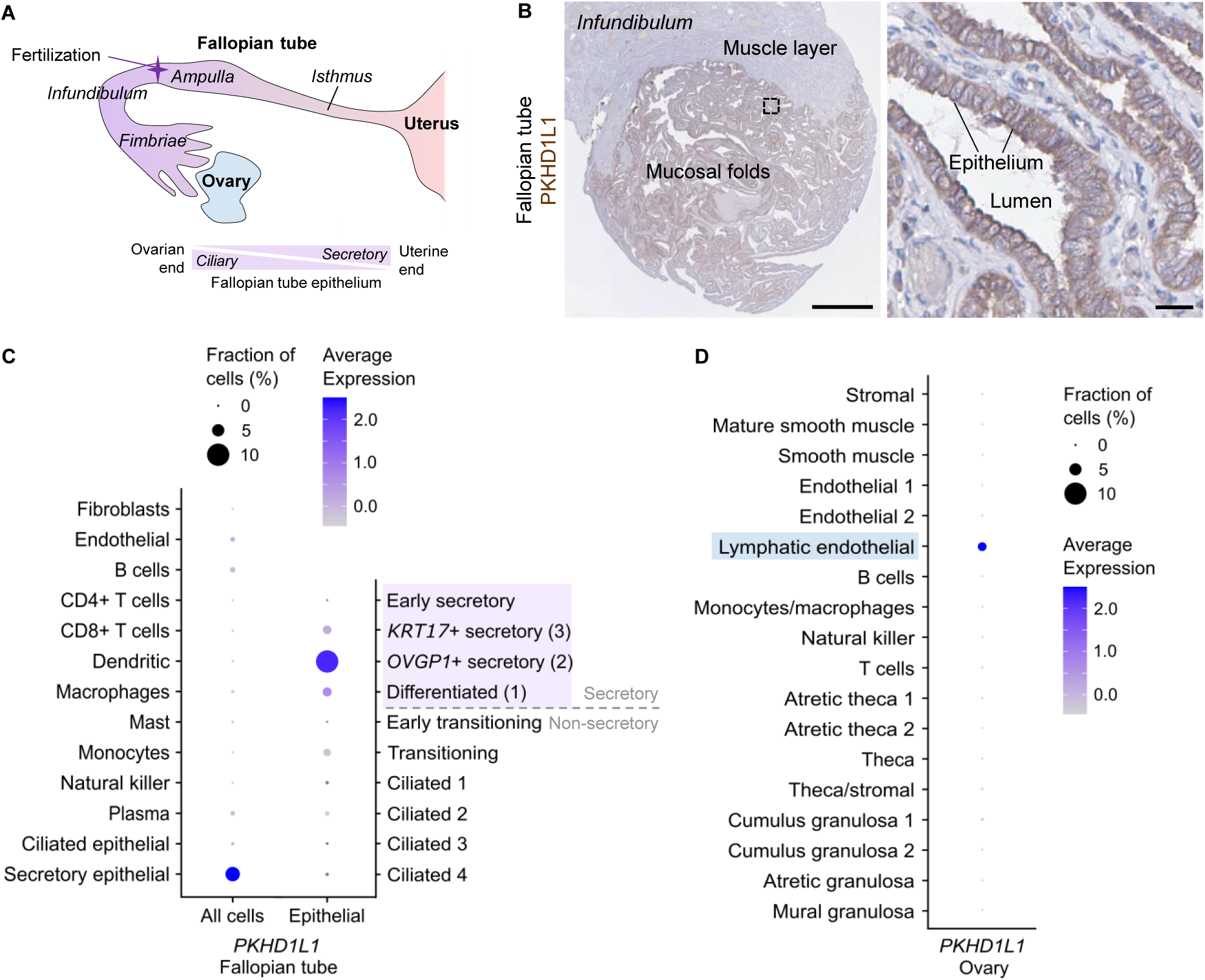
PKHD1L1 is expressed in the human Fallopian tube epithelium. **A**, A graphical presentation of the human Fallopian tube. **B**, Immunohistochemistry of premenopausal Fallopian tube (*n* = 2). Scale bar is 1 mm (left)/25 µm (right). **C**, *PKHD1L1* expression analysis in the human Fallopian tubes; data was generated using the scRNA-seq dataset from ***Dinh et al. 2021***. **D**, scRNA-seq analysis of *PKHD1L1* expression in human ovaries; data was generated using the scRNA-seq dataset from ***Fan et al. 2019***. CD, cluster of differentiation; KRT, keratin; OVGP1, oviductal glycoprotein 1.

### rs17368310 alters PKHD1L1 mRNA splicing

Given that previous GWAS analyses have consistently shown an association between the *PKHD1L1* variant NC_000008.11:g.109459837G>C (hereafter referred to as rs1768310) and female infertility^10,11^, we undertook to specifically assess the functional effects of this variant. First, we investigated how this variant alters mRNA transcription in tissues. Considering that the variant disrupts a canonical GT splice donor site at the junction of exon and intron 47 (**Fig. 3A**), we anticipated that it may lead to either exon skipping or to activation of an alternative (cryptic) donor site within nearby sequence and thus either to loss of sequence or to incorporation of excess sequence into the mRNA^74^. Both deep learning based (SpliceRover^42^, SpliceAI^12^, Pangolin^43^) and RNA-seq based (SpliceVault^44^) splicing prediction software modeled the use of an in-frame cryptic donor site at position +22 in the intron 47 as the most likely alternative splicing event, although other, very rare frameshift-causing exon skipping events and the activation of cryptic donor sites further downstream were also considered possible (**Table S4**). To experimentally verify these prediction results and to study the variant-associated mis-splicing specifically in the uterus, we sequenced a cohort of women based in Northern Finland to find rs17368310 variant carriers (**Fig. 3B**). RNA available from the endometrial biopsies of 5 heterozygous (G/C) donors was then used to synthesize cDNA and examine the most likely affected exon–exon junction site. As expected, primers designed to amplify this region produced one PCR product in the absence of the variant (G/G; **Fig. 3C**). Notably, when amplifying this region in heterozygotes, we detected two PCR products, one corresponding to the reference allele and one ∼20 bp above it (**Fig. 3C and Fig. S2A**). Sanger sequencing revealed that this product resulted from the retention of the first 21 nucleotides of intronic sequence between the exons 47 and 48 (**Fig. 3D**), fundamentally confirming the use of the cryptic donor site as suggested by the prediction tools (**Table S4**). Using forward and reverse primers located further apart, in exons 45 and 51, respectively, we did not detect any transcripts as a plausible result of exon skipping (**Fig. 3E and Fig. S2B**), indicating either their very low to non-existing prevalence (**Table S4**) or immediate nonsense- mediated decay of such transcripts^74^.

**Figure 3.**
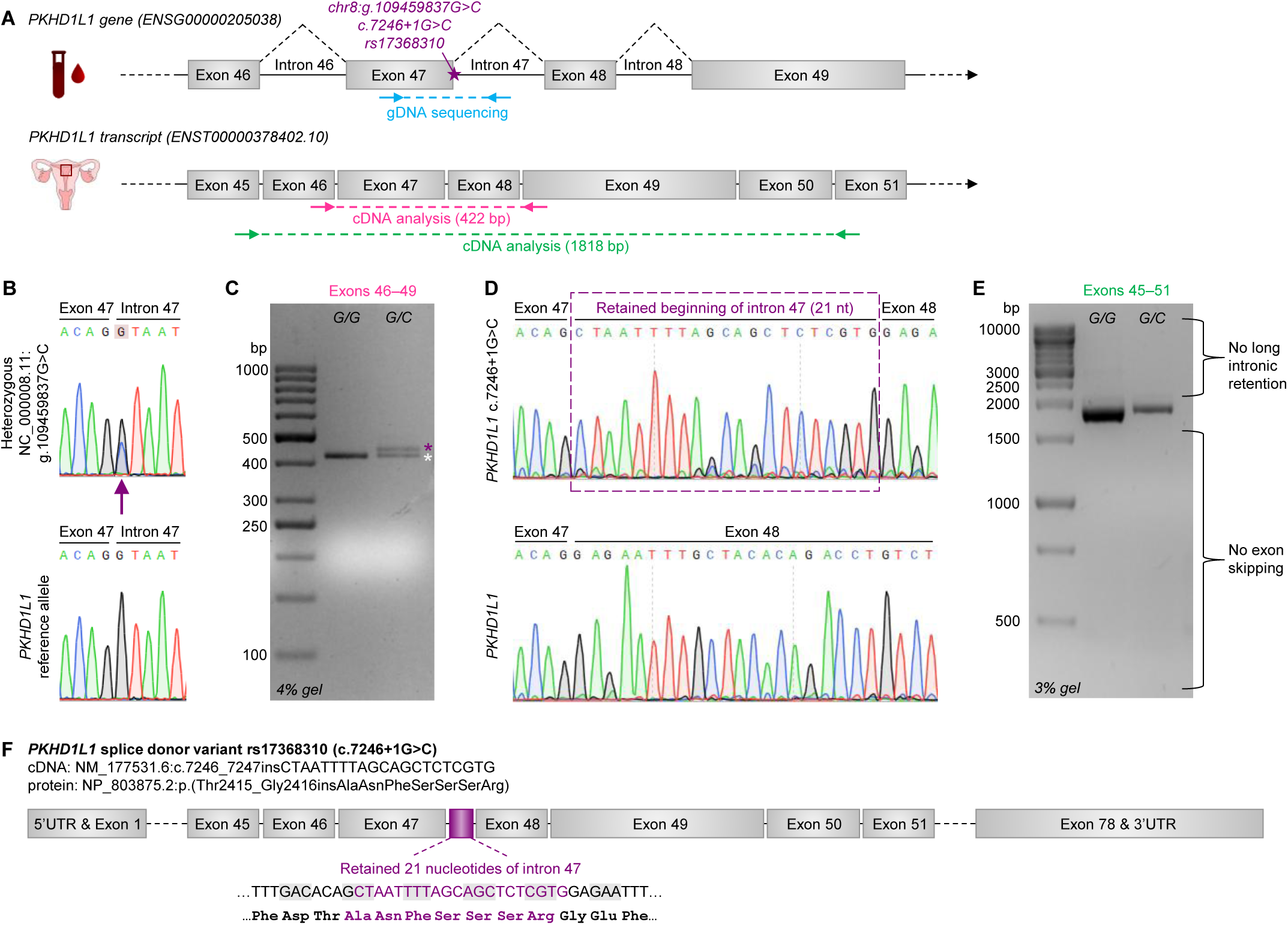
*PKHD1L1* variant rs17368310 disrupts splicing and causes an in-frame intronic sequence insertion. **A**, A schematic representation of *PKHD1L1* genomic DNA (gDNA; upper) and mRNA (lower) displaying the predicted splice donor variant rs17368310 (NC_000008.11:g.109459837G>C; NM_177531.6:c.7246+1G>C) and the study design. **B**, A cohort of women (*n* = 63) was sequenced for the presence of the variant using venous blood–derived gDNA. Heterozygote variant carrier (G/C; *n* = 7) chromatogram shows a double peak of both G and C in the SNP position (purple arrow). **C**, Splicing pattern was analyzed with RT-PCR using endometrial RNA samples of heterozygous donors transcribed into complementary DNA (cDNA) with a primer pair located in exons 46 and 49. White asterisk, PCR product of the reference allele; purple asterisk, variant allele. **D**, Sequencing chromatogram of variant cDNA reveals retention of the first 21 nucleotides of intron 47. *n* = 3 reference and *n* = 5 heterozygote women. **E**, Splicing pattern in the endometrium was further analyzed with RT-PCR with a primer pair located in exons 45 and 51. *n* = 3 reference and *n* = 5 heterozygote women. Note that the 21 bp difference between the reference and variant PCR products of the heterozygote is not visible with this agarose gel percentage and PCR product size. **F**, *PKHD1L1* c.7246+1G>C mRNA contains an in-frame insertion of 21 nucleotides of intronic sequence between exons 47 and 48. The expected amino acid level change, insertion of seven amino acids (purple), is shown. Despite the disruption of the codon for the flanking glycine, there is no change in this amino acid as the final nucleotide of the insertion restores the codon. HGVS Nomenclature–aligning cDNA and protein names based on our data are displayed.

### rs17368310 causes an insertion in a conserved loop of the PKHD1L1 protein

The inclusion of 21 intronic nucleotides in the *PKHD1L1* mRNA causes an in-frame insertion of seven amino acids, ANFSSSR (AlaAsnPheSerSerSerArg), into the translated protein sequence (**Figs. 3F and 4A**). The PKHD1L1 protein (4243 amino acids) is comprised of 14 extracellular Ig-like, plexin, transcription factor (IPT) domains, 10 parallel beta helix 1 (PbH1) repeats, two G8 domains, and two transmembrane protein 2 (TMEM2) homology regions, and it has a very short cytoplasmic tail^14,17,47^ (**Fig. 4A**). PKHD1L1 is also predicted to be proteolytically post-processed^16,46^ (**Fig. 4A**) similarly as shown for the homologue protein PKHD1^75^. The variant-derived insertion affects the first TMEM2-like region of PKHD1L1 (**Fig. 4A**) on an amino acid motif which is well conserved in mammals (**Fig. 4B**, **Table S2**). To understand how this insertion might alter the PKHD1L1 protein structure, we generated AlphaFold^50^ models of the native first TMEM2-like region and of the one containing the insertion, mimicking the rs17368310 variant. The folded core structure highly resembles that of TMEM2, with some differences in the loop regions and additional domains (**Fig. S3**)^55^. In PKHD1L1, this region contains a central β-helix, which begins with an N-terminal G8 domain connected to a larger folded domain (**Fig. 4C**). After the first turns of the β-helix, there is an inserted β-sandwich domain and, one strand after the β-sandwich, a 43-residue loop within the β- helix (residues Ser2397–Gly2439; **Fig. 4C**). In the predicted PKHD1L1 structure, this loop, termed Loop 1, is packed against the main β-helix with high confidence (**Fig. 4D, Fig. S4**). The tip of Loop 1 is located on the surface of the structure model together with two other loops (Loop 2/residues Phe2545–Ala2561 and Loop 3/residues Thr2757–Gly2769). The variant-derived insertion locates between Thr2415 and Gly2416 before the tip of Loop 1, and it causes apparent conformational changes within the loop according to the AlphaFold prediction (**Fig. 4E, Fig. S4**). Consequently, the modification is likely to cause alterations in the potentially functionally important surface formed by the three loops, while the overall structure of Loop 1 outside the immediate tip region is well preserved. To expand the mammalian sequence conservation analysis (**Fig. 4B**), we generated a multiple sequence alignment of 87 PKHD1L1 proteins in different vertebrates and mapped the conservation on the surface of the human PKHD1L1 structural model. This showed that Loops 1 and 2 form a surface that is highly conserved across a diverse range of vertebrate species, while other parts of this TMEM2-like region are somewhat less conserved **(Fig. S5, Table S2**). The potential functional role of Loop 1 is therefore supported both by its apparent non-structural role and high degree of conservation in different species.

**Figure 4.**
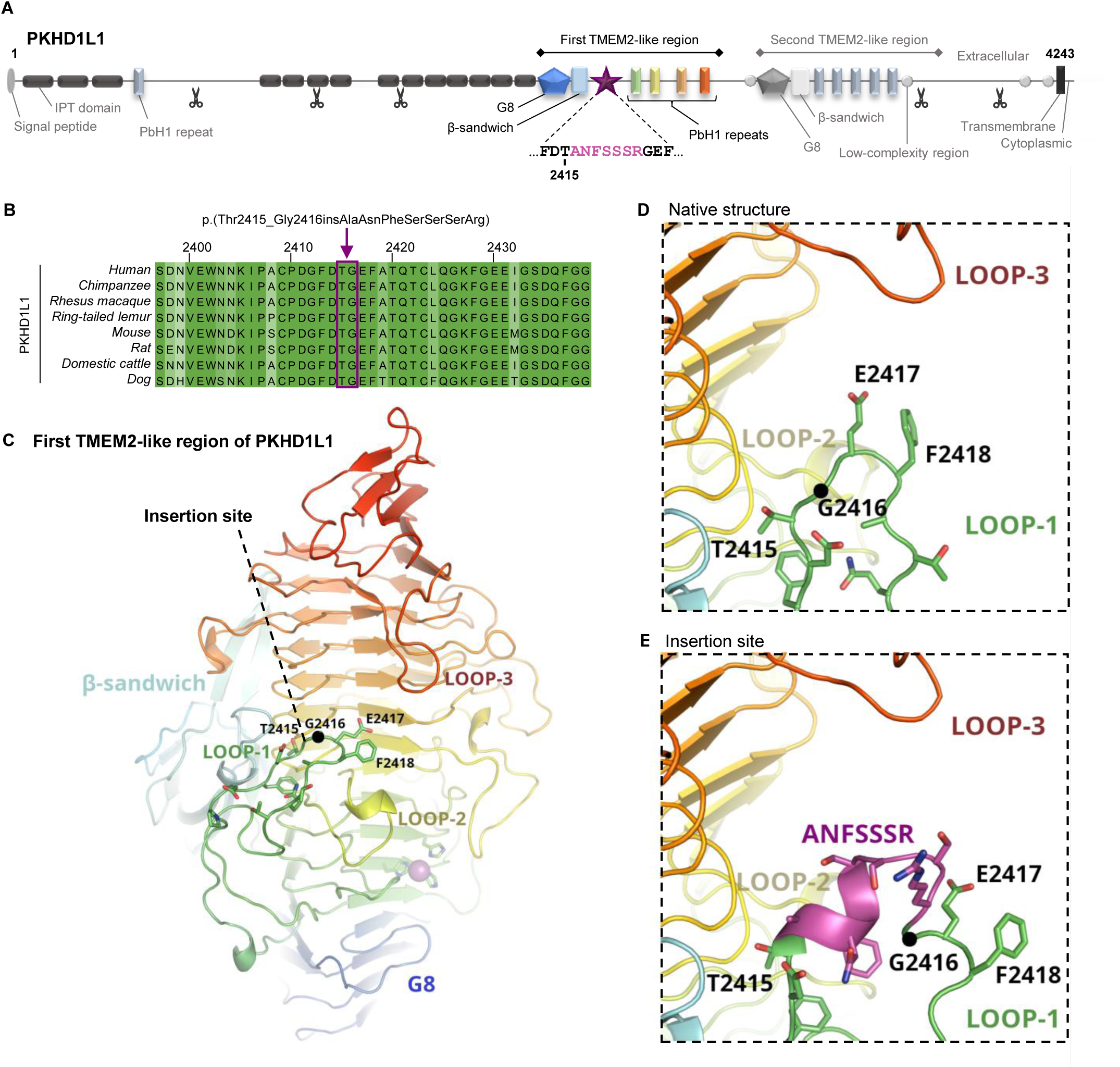
Modeling of the structural alterations in PKHD1L1 protein as a result of *PKHD1L1* variant rs17368310. **A**, Human PKHD1L1 protein domain map based on database analysis. Scissors mark predicted general proprotein convertase cleavage sites on arginine residues 772, 1250, 1654, 3567, and 3987. The variant-derived splicing change is predicted to result in an addition of seven amino acids within the first TMEM2-like region (p.Thr2415_Gly2416insAlaAsnPheSerSerSerArg; magenta star). **B**, PKHD1L1 protein sequence alignment in common mammals. The red arrow and box mark the site of the predicted insertion within Loop 1 depicted in panel C. Amino acid numbering is based on the human reference sequence (including the signal peptide). The higher the color intensity, the more similar the residue is across the shown species. **C**, AlphaFold model of the first PKHD1L1 TMEM2-like region. Loop 1 containing the insertion site and two other loops (Loops 2 and 3) forming a potentially important functional surface are labeled. Conserved histidine residues form a predicted binding site for a metal ion shown as a magenta sphere. **D**, Close-up view of Loop 1 in the native protein. **E**, Close-up view of Loop 1 with the variant-derived ANFSSSR sequence insertion (magenta). IPT, Ig-like, plexin, transcription factor; PbH1, parallel beta helix repeats; TMEM2, transmembrane protein 2.

Post-translational modifications (PTMs) of the insertion site might further exacerbate the structural changes within the mutated protein domain and hinder its proper function. We noted how the insertion ANFSSSR creates multiple potential novel PTM sites (**Table S5**), including a consensus sequence for N-glycosylation (NXS, where X ≠ Pro)^58^ and three serines, which can be targets for either O- glycosylation^59^ or phosphorylation^60,61^. Despite the general disagreement between the prediction software on the likelihood of each individual PTM (**Table S5**), the multitude of potential PTM sites suggests that PTMs may impact the insertion and, thereby, protein function and stability.

### rs17368310 associates with female infertility and related traits in the Finnish and Mexican populations

Genetic variants at the *PKHD1L1* locus, including the likely causal variant rs17368310, have been reported to be associated with female infertility under both additive and recessive models^10,11^. Additionally, women carrying two copies of the alternative rs17368310 allele (*i.e*., C/C homozygotes) were previously reported to have a slightly reduced child count in the Finnish population^10^. Here, using data from the FinnGen project^62^, we investigated whether the *PKHD1L1* locus is similarly associated with an even more specific infertility-related phenotype, focusing on women medically treated for infertility with oral ovulation induction medication (clomifene), follicle stimulating hormone analogues, and/or gonadotropins (**Table S3**). In addition, we analyzed the association with the mean number of children utilizing data extracted from the Finnish Population and Medical Birth Registries. Single-nucleotide polymorphisms (SNPs) at the *PKHD1L1* locus were associated with medical treatment for infertility under the recessive model (case/control *n* = 9,381/272,683; **Fig. 5A, Table S6**). The association was less significant under the additive model, indicating that variants at this locus are likely to act recessively, consistent with previous findings^10^. The lead SNP at the locus was an intronic *PKHD1L1* SNP, rs4735131, which is highly correlated (linkage disequilibrium, *r*^2^ = 0.94) with rs17368310; therefore, these SNPs represent the same association signal. Of all associated variants, rs17368310 was the only variant with direct predicted functional consequences (**Table S6**). The effect estimate of rs17368310 (odds ratio 2.07, 95%CI 1.63–2.64 for the C/C homozygotes, *p* = 4 x 10^-9^; **Table S6**) for medical treatment for infertility corresponded to that previously reported for female infertility in FinnGen^10^. Of all women who had reached the age of 45 years at FinnGen, 15.4% did not have children (**Table S7**), the mean number of children being 1.94 (**Table S8**). A larger proportion of C/C homozygotes at rs17368310 had been medically treated for infertility compared to the other genotypes (5.46% vs. 2.95%); a larger proportion also did not have children compared to women with the other genotypes (19.0% vs 15.4% C/C vs C/G and G/G, *p* = 0.004; **Table S7**). Consistently, the mean number of children was lower in individuals with the C/C genotype (1.78 vs 1.94, *p* = 0.003) (**Fig. 5B, Table S8**). Interestingly, we noted that this difference was even more evident among the women who had received medical treatment for infertility (proportion not having children 44.9% vs 23.2% C/C vs other genotypes, *p* = 0.0007; mean number of children 0.94 vs. 1.53, *p* = 0.0002) (**Fig. 5C, Tables S7 and S8**), indicating that the infertility phenotype of rs17368310 C/C homozygote women in the Finnish population was not rescued by the medical infertility treatments.

**Figure 5.**
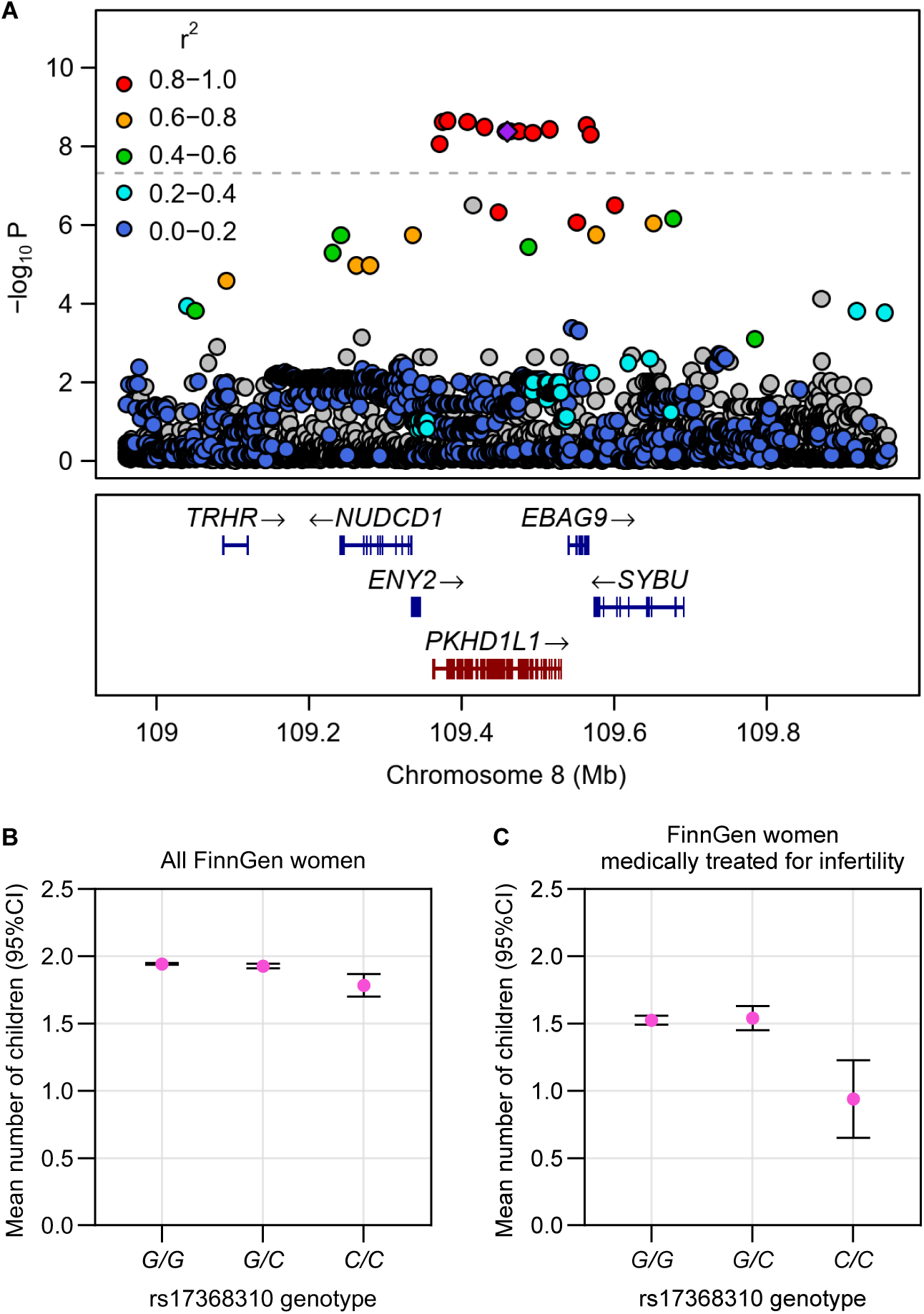
Association of rs17368310 with medical treatment for infertility and with the number of children. A, In the regional association plot, each data point is a SNP; position on chromosome 8 is indicated on the X axis, and the negative logarithm of p value is shown on the Y axis. Linkage disequilibrium (r^2^) is indicated with varying colors with respect to rs17368310 which is shown as a purple diamond, and genes are shown below. **B**, Mean number of children in all FinnGen women who had reached the age of 45 years according to the rs17368310 genotype. **C**, Mean number of children in the FinnGen women medically treated for infertility who had reached the age of 45 years according to the rs17368310 genotype. Medical treatment for infertility included ovulation-inducing drugs, follicle stimulating hormone, and/or gonadotropin. Data in **B** and **C** is presented as mean±95% confidence interval (CI).

We further investigated the allele frequencies of rs17368310 in different populations and noted that it has an average global allele frequency of 5.1%, being more than 20% in the admixed American populations, 6.8% in the Finnish population, ∼5% in the non-Finnish Europeans and South Asians, and less than 2% in the African Americans and East Asians (gnomAD^76^ v4.1.0, https://gnomad.broadinstitute.org/). Given the reported markedly higher allele frequency in the admixed American population, we sought to replicate our findings in the Mexico City Prospective Study (MCPS) cohort representing the Central American population, with a notably higher rs17368310 allele frequency (31.1% in the MCPS cohort, **Table S9**) compared to most other populations. As information on the number of children was not available from the MCPS cohort, we used questionnaire data for the number of pregnancies as a surrogate phenotype. We detected that the self-reported number of pregnancies was lower in rs17368310 C/C homozygotes compared to other genotypes (mean number of pregnancies 4.73 vs 4.83, **Table S9**), consistent with the association of the C/C genotype with the number of children in the FinnGen data. Furthermore, rs17368310 associated with the number of pregnancies, and this association was clearly more pronounced under the recessive model (*p* = 6 × 10^-13^) compared to the additive model (*p* = 7 × 10^-9^; **Table S9**).

To investigate if rs17368310 is associated with other phenotypes, we investigated previously reported associations of this SNP from the GWAS Catalog^65^ and FinnGen endpoints^62^. There were no associations for this SNP in the GWAS Catalog, and only three associated SNPs were indicated when the whole *PKHD1L1* region was queried. As expected, one of these was rs9643050 (in total LD with rs17368310) associated with female infertility in the previous GWAS for infertility^11^; the two other associations were with height^77^ and with circulating levels of CES1 (liver carboxylesterase 1)^78^ for SNPs (rs10100944, rs1457282) non-correlated with the infertility-associated SNPs. We only detected the following two associations in FinnGen: Female infertility (*p* = 2.9 × 10^-13^) and Female infertility, cervical, vaginal, other or unspecified origin (*p* = 1.4 × 10^-13^).

## Discussion

Modern genome analysis methods have rapidly increased our understanding of the association of designated genetic loci and variants with diseases. However, the true impact of these gene variants on the molecular and cellular functions often remains enigmatic due to the lack of follow-up studies. Herein, we have examined one such association and dissected the connection of *PKHD1L1* with female infertility. Although the association of *PKHD1L1* with female infertility had been suggested in earlier studies^9–11^, the observation was somewhat overshadowed by the adjacent coding gene, *EBAG9*, which is also an estrogen-responsive gene and was proposed to underlie the association^9,11^. In this study, we demonstrated that *PKHD1L1* is the plausible causal gene at this locus. We showed that *PKHD1L1* expression is dynamically regulated across the menstrual cycle in the female reproductive tract, the predicted *PKHD1L1* splice donor variant rs17368310 indeed disarrays *PKHD1L1* mRNA splicing in the endometrial tissue, and the variant is likely to affect normal protein function based on structural biocomputational analysis. We observed an increased frequency of rs17368310 homozygotes among women medically treated for infertility, accompanied by reduced child count. Notably, the infertility treatments did not rescue the genotype effect that was present also in the treated women. In addition to these data derived from the Finnish population, data from a large Mexican-based population study showed fewer pregnancies among women homozygous for rs17368310. Altogether, our study provides compelling evidence on the role *PKHD1L1* plays in supporting female fertility.

To understand more profoundly the time window for the action of *PKHD1L1*, we comprehensively mapped the spatiotemporal expression pattern of PKHD1L1 in the female reproductive tract both by using unique in-house tissue materials and by collecting together the previously scattered evidence from large-scale datasets. All the generated data provide broad evidence on the epithelium-enriched expression of *PKHD1L1* predominantly in the pre-receptive endometrium and in a specific subset of *OVGP1*+ Fallopian tube secretory cells known to emerge just before ovulation^32^. These results are further supported by other recent transcriptomic studies^22,28^. This specific timing and location of the peak expression suggest that PKHD1L1 may be needed to prime the endometrium for decidualization and the window of implantation, and that it may play a role in gamete or zygote preparation. In turn, the drop in PKHD1L1 expression levels in mid- and late secretory endometrium implies its expression needs to be suppressed during embryo implantation. Along with this, MRGDv2 data showed that the decidua during early pregnancy and placenta do not express PKHD1L1, indicating that defects in the development of the implanted embryo and placentation are unlikely to explain the lower child count in women homozygous for rs17368310.

Previous studies have reported PKHD1L1 expression specifically in the ciliated cells of the ear, heart, and bile duct^15,16,30,79^. Yet, our data clearly indicate that in the female reproductive tract, secretory epithelial cells are the main epithelial cell type expressing PKHD1L1, while the ciliary epithelium shows lesser expression, suggesting that PKHD1L1 has functions beyond the ciliary machinery. Similarly to some other large glycoproteins^73,80^, PKHD1L1 could be secreted to the uterine and/or Fallopian tube fluid, since most of its extracellular part is predicted to be proteolytically cleaved off from the cell membrane. The antibody we used for immunohistochemistry targets a region so close to the transmembrane domain (aa 4105–4186) that it would remain bound to the membrane even if shedding were to occur, however, proteomics data show the presence of PKHD1L1 protein in the uterine fluid^81^, further suggesting its secretion.

We experimentally confirmed the effect of rs17368310 on *PKHD1L1* mRNA splicing in the endometrium. The SNP eliminates the normal splice donor site of exon 47, which induces the use of a cryptic donor site 21 bp into intron 47. Given that this preserves frame reading, the faulty mRNA is likely, as we showed for the heterozygote variant carriers, to escape nonsense-mediated decay. In line with this, the relatively mild phenotype of the variant carriers—fertility is slightly declined but not fully abolished—does not suggest a complete loss-of-function of the protein but rather only partially hampered functionality. This aligns with our AlphaFold modeling results and the extensive size and complexity of the protein, rendering it likely that other domains outside the impacted region are still structurally intact and functional. Nevertheless, while the overall variant protein structure is likely preserved, our structural modeling showed that local changes in the affected loop may compromise its interaction with the other nearby loops, thereby disturbing correct function. G8 domain and PbH1 repeats, which are located one after the other with the loop sequences in PKHD1L1 protein, often coexist in the same proteins and are involved in carbohydrate binding, scaffolding, and/or catalysis^47,75^. Interestingly, PKHD1L1 homologue protein TMEM2 seems to play a part in hyaluronic acid metabolism^55,82–84^, and the regulation of the molecular weight of hyaluronic acid has been connected with fertilization success^85^. However, as the functional roles of TMEM2 and another PKHD1L1 homologue, PKHD1, are still heavily debated^55,75,82,83,86^ and our structural comparison revealed not only similarities but also differences in the loop structures between PKHD1L1 and TMEM2, establishing this connection for PKHD1L1 will require substantial further studies.

Our analyses showed that although women homozygous for the rs17368310 variant can conceive naturally, they are seeking for medical treatments for infertility more often than the other genotypes. Critically, the infertility phenotype of these women is not rescued by the common repertory of medical infertility treatments, often also referred to as assisted reproductive technologies (ART). Treatments specifically included in the used FinnGen data are either solely aimed at stimulating the ovaries (clomifene, follicle stimulating hormone) or involve a more complex hormonal treatment path (follicle stimulating hormone, gonadotropins) with embryo transfer and luteal support as in the case of *in vitro* fertilization (IVF). As we observed that PKHD1L1 is not significantly expressed in the ovaries, it is tempting to speculate that mere ovulation induction, the usual first-line treatment choice combined with intrauterine insemination^87^, may therefore not be very effective in treating homozygous patients and does not solve the PKHD1L1-related dysfunction affecting the Fallopian tube and endometrial lining. Furthermore, while IVF treatment bypasses the Fallopian tubes, no infertility treatment can fully overcome the dysfunctionality of the endometrial component. Substantial research has been carried out on the genetic variants impacting human ovarian function and embryo quality^4,5^, and, rather surprisingly, less focus has been given to the environment where the embryo develops—the Fallopian tubes and the uterus^8^. Thus, our study adds significantly to existing efforts and highlights the critical importance of investigating also the detailed molecular underpinnings of the endometrium and Fallopian tubes to fully understand pregnancy establishment in humans. As for the patient perspective, naming the potential defect may help the psychological burden of unexplained infertility but also pave the way for improved diagnostics and novel therapeutic approaches.

Limited availability of human tissues and related datasets prohibited us from assessing *PKHD1L1* expression in the ovarian cortex containing surface epithelial cells and in the Fallopian tube isthmus. Furthermore, all available Fallopian tube sample sets, if any menstrual cycle timing was attained, use *OVGP1* expression as a proxy for menstrual cycle phase determination^32,33,69,70^, thereby making it difficult to directly date the peak of *PKHD1L1* expression, which, nevertheless, likely is a rather transient event given the reported estrogen-dependency of *OVGP1*+/*PKHD1L1*+ secretory cells^32,33^ and the short peak of high estrogen levels around ovulation. Further dissection of the individual medical treatments for infertility among the C/C homozygotes was not feasible due to low case count. Finally, analysis of the Mexican population was based on the self-reported number of pregnancies as the surrogate infertility phenotype; a complementing study using data on the number of born children in high rs17368310 allele frequency populations remains to be established.

The FinnGen data on the Finnish population has enabled the identification of the clinical effects that rs17368310 has on female fertility, and importantly, our further analysis of the MCPS data on the Mexican population serves as an additional confirmation of its trans-ethnic implications. The persistence of fertility-reducing genetic variants in the population should, in general, be low; surprisingly, the *PKHD1L1* locus does not seem to be affected by negative selection in humans (https://reich-ages.rc.hms.harvard.edu/#/), possibly pointing to the presence of some balancing positive selection that yet remains unknown. Using the United Nations information on population size in 2025 and the MCPS Mexican allele frequency 0.311, there are over 6.3 million homozygous women in Mexico, and, using the 1000 Genomes Project Peruvian allele frequency 0.459, there are over 3.6 million homozygous women in Peru, to name a few of the most highly impacted populations. Consequently, we can estimate that rs17368310 affects the fertility of millions of women across the world, and particularly those of Latin American ancestry. Our study hence provides globally highly relevant knowledge on the impact of the *PKHD1L1* gene and its variant rs17368310 in controlling female fertility.

## Supporting information

Kapiainen et al. Supplementary Materials

## Data Availability

All data produced in the present work are contained in the manuscript

## Acknowledgements

We gratefully acknowledge all tissue sample donors and medical personnel contributing to this research. We thank Biocenter Oulu Sequencing Center, University of Oulu, for providing expert services and The Department of Pathology & Immunology at Baylor College of Medicine for access to the Sectra Digital Pathology Services. We also thank Dr. Audrey Savolainen, Felix Bächtle, Maarit Haarala, and Célia Tebbakh for technical assistance and Prof. Lloyd Ruddock, University of Oulu, for discussions and critical revision of the manuscript draft.

This work was supported by funding from the University of Oulu and the Research Council of Finland Profi6 Fibrobesity program 336449 (E.K., M.K.K., and R.P.-H.); Biocenter Oulu Emerging Project (R.P.-H.); the Sigrid Jusélius Foundation (T.T.P. and R.P.-H.); Fulbright Finland Foundation (E.K.); Novo Nordisk Foundation (T.T.P.); and GeneCellNano Flagship of the Research Council of Finland (P.B.P.).

We want to acknowledge the participants and investigators of the FinnGen study. A full FinnGen consortium author list can be found in **Table S10**. The FinnGen project is funded by two grants from Business Finland (HUS 4685/31/2016 and UH 4386/31/2016) and the following industry partners: AbbVie Inc., Alnylam Pharmaceuticals, Inc., AstraZeneca UK Ltd, Bayer AG, Biogen MA Inc., Boehringer Ingelheim International GmbH, Bristol Myers Squibb Inc. (and Celgene Corporation & Celgene International II Sàrl), Genentech Inc., GlaxoSmithKline Intellectual Property Development Ltd., Johnson&Johnson Innovative Medicine Inc., Maze Therapeutics Inc., Merck Sharp & Dohme LCC, Novartis AG, Pfizer Inc. and Sanofi US Services Inc. Following biobanks are acknowledged for delivering biobank samples to FinnGen: Auria Biobank (www.auria.fi/biopankki), THL Biobank (www.thl.fi/biobank), Helsinki Biobank (www.helsinginbiopankki.fi), Biobank Borealis of Northern Finland (https://www.ppshp.fi/Tutkimus-ja-opetus/Biopankki/Pages/Biobank-Borealis-briefly-in-English.aspx), Finnish Clinical Biobank Tampere (www.tays.fi/en-US/Research_and_development/Finnish_Clinical_Biobank_Tampere), Biobank of Eastern Finland (www.ita-suomenbiopankki.fi/en), Central Finland Biobank (www.ksshp.fi/fi-FI/Potilaalle/Biopankki), Finnish Red Cross Blood Service Biobank (www.veripalvelu.fi/verenluovutus/biopankkitoiminta), Terveystalo Biobank (www.terveystalo.com/fi/Yritystietoa/Terveystalo-Biopankki/Biopankki/) and Arctic Biobank (https://www.oulu.fi/en/university/faculties-and-units/faculty-medicine/northern-finland-birth-cohorts-and-arctic-biobank). All Finnish Biobanks are members of BBMRI.fi infrastructure (https://www.bbmri-eric.eu/national-nodes/finland/). Finnish Biobank Cooperative – FINBB (https://finbb.fi/) is the coordinator of BBMRI-ERIC operations in Finland.

Part of this research has been conducted using Mexico City Prospective Study (MCPS) data under Application Number 2025-082. The MCPS (https://www.ctsu.ox.ac.uk/research/mcps) is a long- standing scientific collaboration between researchers at the National Autonomous University of Mexico, Mexico, and the University of Oxford, UK, and has received funding from the Mexican Health Ministry, the National Council of Science and Technology for Mexico, Wellcome, Cancer Research UK, the British Heart Foundation, Kidney Research UK, and the UK Medical Research Council.

## Author contributions

Conceptualization: EK, JK, RPH

Data Curation: EK, MKK, PBP, ET, DAR, Lle

Formal Analysis: EK, MKK, PBP, ET, DAR, Lle

Funding Acquisition: EK, MKK, PBP, TTP, RPH

Investigation: EK, MKK, PBP, SEP, ET, DAR, Lle

Methodology: -

Project Administration: JK, RPH

Resources: MKK, RKA, US, LLu, MM, JMT, JB, JAD, PKM, RTC, LCC, RPM, KP, DM, TTP, JK

Software: -

Supervision: DM, TTP, JK, RPH Validation: EK, MKK

Visualization: EK, MKK, PBP, LLe

Writing—Original Draft: EK, MKK, LLe, JK, RPH Writing—Review & Editing: All authors reviewed the work.

## Data and code availability

Data and materials are available upon reasonable request from the corresponding author whenever possible but may be subject to stringent restrictions due to legal and ethical reasons or require a completed data or materials transfer agreement. Some of the analyses in this paper are based on published datasets and analysis tools that are available via the referenced studies and websites. The Finnish biobank data can be accessed through the Fingenious services (https://site.fingenious.fi/en/) managed by FINBB. Data from the Mexico City Prospective Study are available to bona fide researchers. The MCPS Data and Sample Sharing policy can be viewed (in English or Spanish) on the study webpage (https://www.ctsu.ox.ac.uk/research/mcps), and the questions utilized in the study, along with the available study data, can be accessed and reviewed through the study’s Data Showcase (https://datashare.ndph.ox.ac.uk/mexico/).

