## Supplementary material for "*PKHD1L1* affects fertility in women": Kapiainen et al. Supplementary Materials

### Figure S1

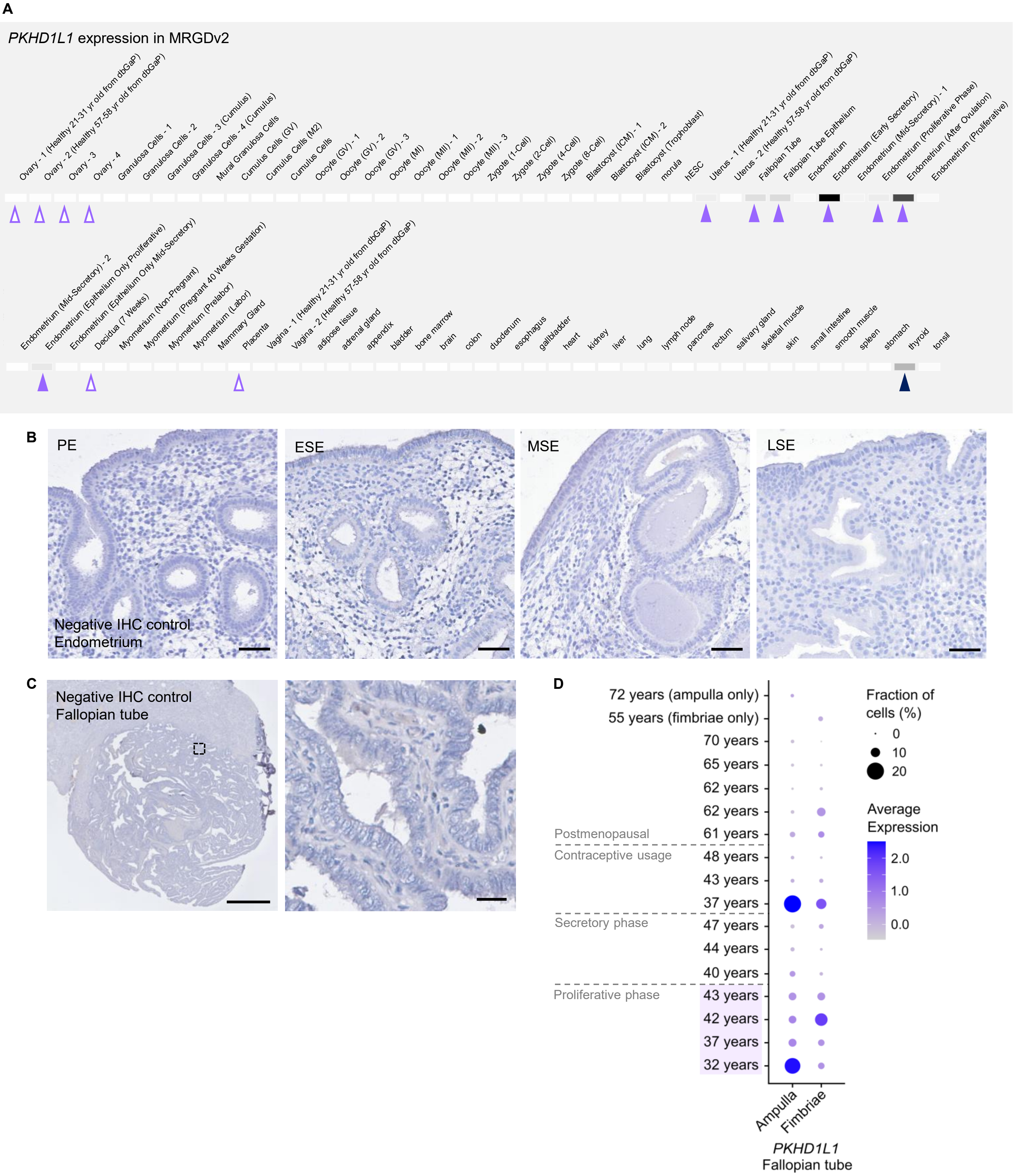

**Figure S1. Supporting data for PKHD1L1 expression analyses. A**, Analysis of *PKHD1L1* expression in both the female reproductive tract and non-reproductive tissues using the Mammalian Reproductive Genetics Database v2 (https://orit.research.bcm.edu/MRGDv2). The darker the bar, the higher *PKHD1L1* expression is in the corresponding tissue. Note especially high the *PKHD1L1* expression in Fallopian tubes and in proliferative/early secretory endometrium (lilac arrowheads). Ovaries, decidua, and placenta do not show *PKHD1L1* expression (empty arrowheads). Of the non-reproductive tissues, the thyroid shows high expression (navy arrowhead). **B–C,** Negative (no primary antibody) controls of the PKHD1L1 IHC stainings shown in **Figure 1C** for endometrium (**B**) and in **Figure 2B** for Fallopian tube (**C**). Scale bar is 50 µm in **B** and 1 mm (left) and 25 µm (right) in **C**. Counterstaining with hematoxylin. ESE, early secretory endometrium; MSE, mid-secretory endometrium; LSE, late secretory endometrium; PE, proliferative endometrium. **D,** scRNA-seq analysis of *PKHD1L1* expression in the Fallopian tubes generated using the dataset of ***Weigert et al. 2025***.

### Figure S2

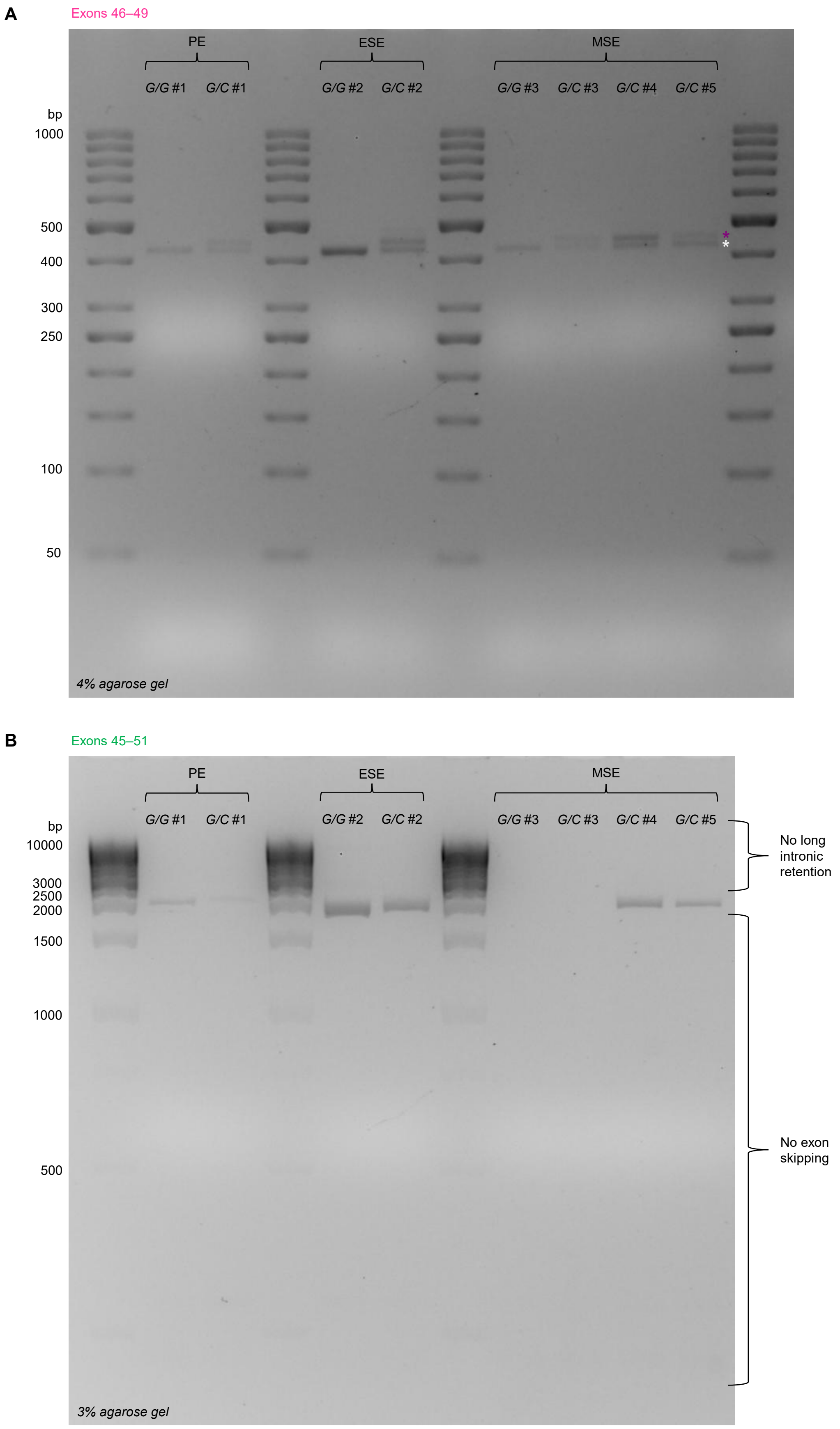

**Figure S2. rs17368310 splicing pattern analysis.** Splicing pattern of *PKHD1L1* variant rs17368310 was analyzed with RT-PCR using endometrial RNA samples with primer pairs locating in exons 46 and 49 (**A**) or in exons 45 and 51 (**B**), depicted in **Figure 3A**. $n$ = 3 reference (G/G) and $n$ = 5 heterozygous (G/C) women. All analyzed heterozygous samples are shown here, and one example in **Figures 3C and 3E**. ESE, early secretory endometrium; MSE, mid-secretory endometrium; PE, proliferative endometrium. White asterisk, PCR product of the reference allele; purple asterisk, variant allele. The gel images are unadjusted for any enhancement.

### Figure S3

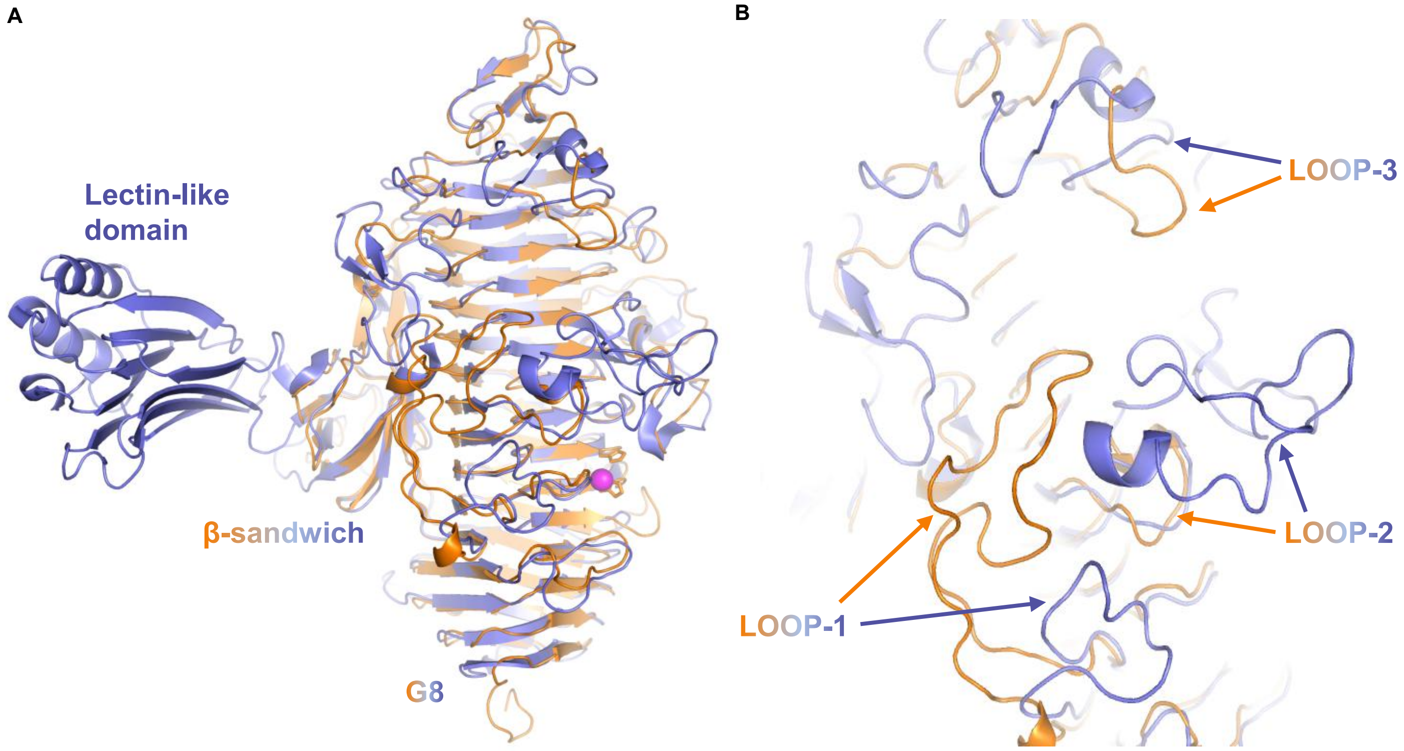

**Figure S3. Superimposition of the PKHD1L1 AlphaFold model with the crystal structure of TMEM2. A**, The crystal structure of residues Asn112–Ser990 of TMEM2 ectodomain (PDB ID 8C6I) is shown in blue and the AlphaFold model of the first TMEM2-like region of PKHD1L1 in orange. $Ni^{2+}$ of TMEM2 is shown as a magenta sphere. PKHD1L1 lacks the lectin-like domain found in TMEM2. **B**, A zoomed-in view of the surface containing the loop structures. Loop 1 has 43 residues in PKHD1L1 and 20 residues in TMEM2.

### Figure S4

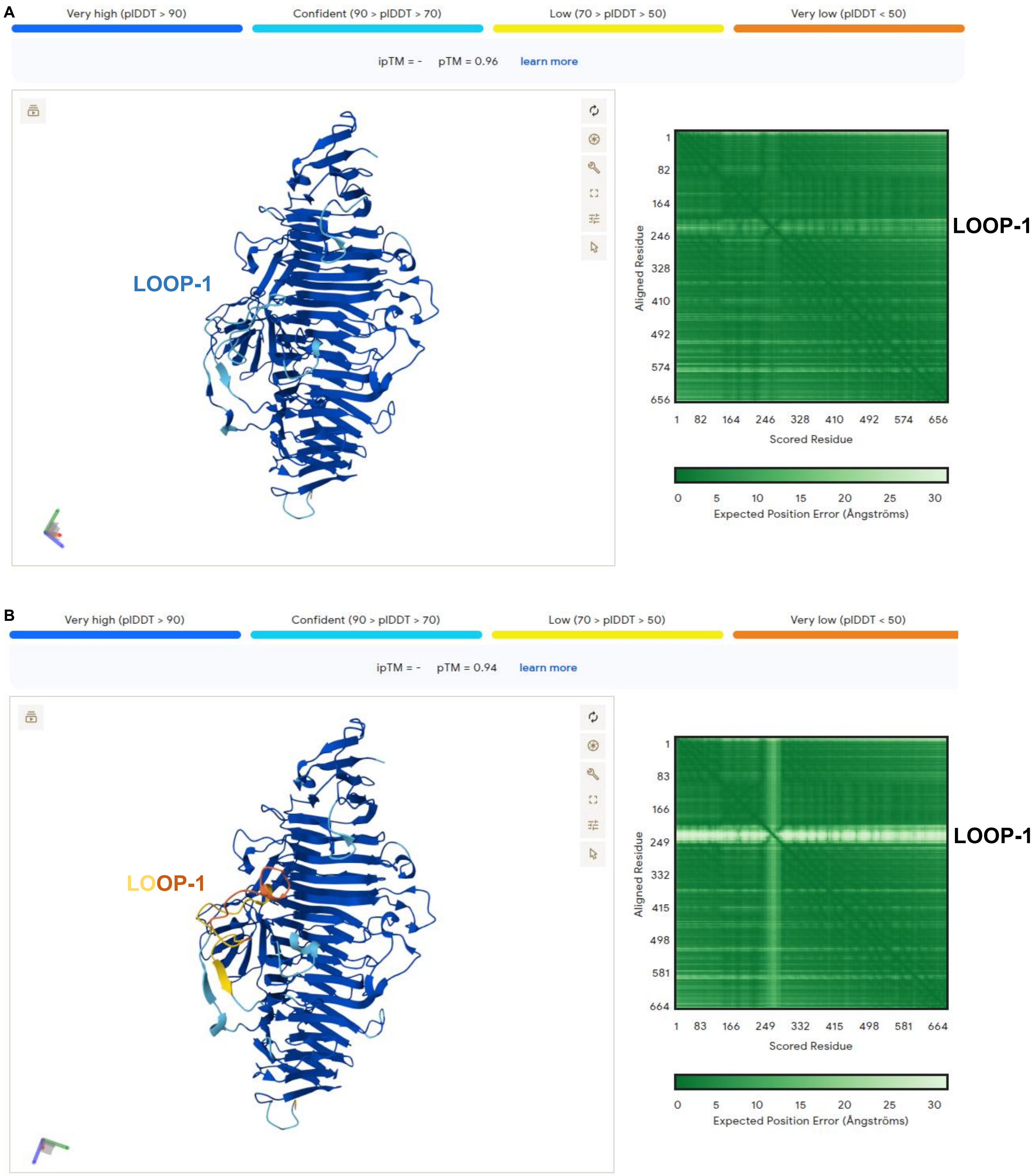

**Figure S4. Supporting data for modeling of the structural alterations in PKHD1L1 protein as a result of PKHD1L1 variant rs17368310.** AlphaFold model for the native (**A**) and variant (**B**) PKHD1L1 protein fragments shown in **Figure 4C–E**. **A,** The model for native PKHD1L1 is of high confidence throughout, and the positional errors are small. **B**, Variant protein has low pIDDT scores and large position error for the loop region following the insertion (Loop 1), as highlighted with the yellow and orange colors. ipTM, interface predicted template modeling score; pLDDT, predicted local distance difference test; pTM, predicted template modeling score.

### Figure S5

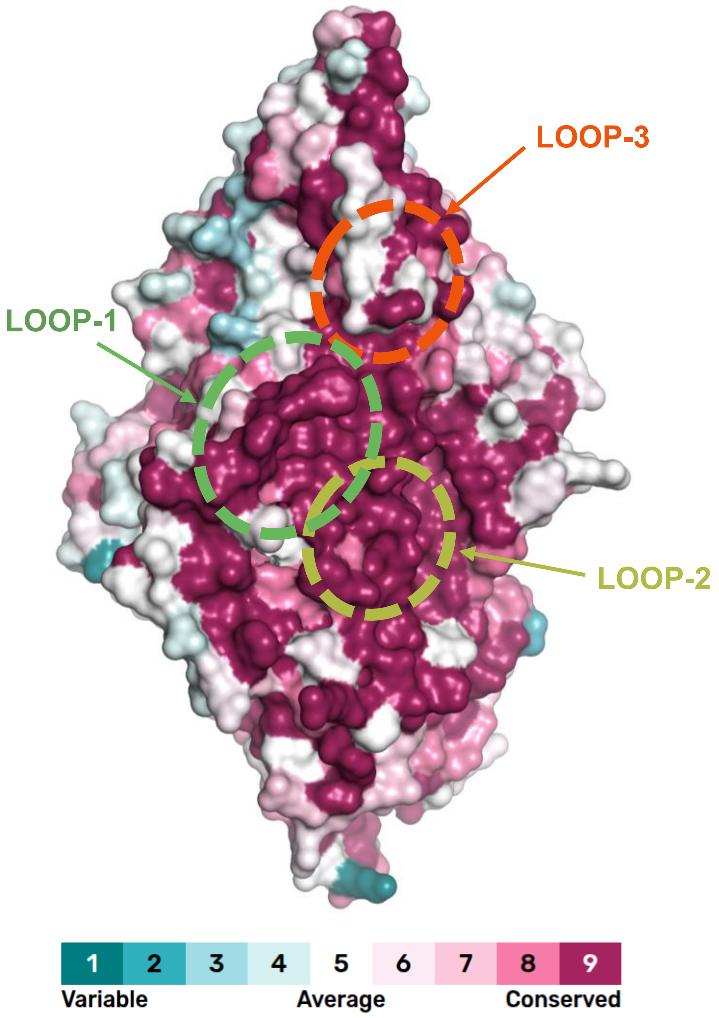

**Figure S5. Structural conservation analysis of the first PHKD1L1 TMEM2-like region.** The conservation was mapped to the structural AlphaFold model using multiple sequence alignment of 87 different vertebrate species (**Table S2**). The three loops on the surface of the β-helix are circled.

#### Table S1. Primers used in this study.

| Primer | Sequence (5' — 3') | Description |
|---|---|---|
| *PKHD1L1*_For1 | GAGCAGAAGTTGGAATTCTTACAA | FFPE gDNA sequencing (exon 47 to intron 47); 141 bp product |
| *PKHD1L1*_Rev1 | TGGCATGATTATACCAACCACG | |
| *PKHD1L1*_For2 | ATCTGTGTCTGCTGATGGCA | Blood gDNA sequencing (exon 47 to intron 47); 404 bp product |
| *PKHD1L1*_Rev2 | ACTATCAGCATTTCCAAATTTCCAC | |
| *PKHD1L1*_For3 | GTAATTGCAAGCACAGGACACAG | cDNA sequencing (exons 46 to 49); 422 bp product |
| *PKHD1L1*_Rev3 | GCCTGGCCAGCATGGAATAC | |
| *PKHD1L1*_For4 | AGGGAATCCTGGATCTGCACG | cDNA sequencing (exons 45 to 51); 1818 bp product |
| *PKHD1L1*_Rev4 | GAATCAGAGCCATCCATCCAGAC | |
| *PKHD1L1*_For5 | TCAGAGTTCTAGTTGGAGGTGAA | RT-qPCR (exon 11/12 junction to exon 13); 162 bp product; Primer Bank ID 126116588c3 |
| *PKHD1L1*_Rev5 | CCAAACGTATTGGACGGCTATT | |
| *GAPDH*_For | GCTCATTTCCTGGTATGACAACG | RT-qPCR (exon 5/6 junction to exon 6); 187 bp product |
| *GAPDH*_Rev | GGGGAGATTCAGTGTGGTGG | |

#### Table S2. PKHD1L1 amino acid sequences used for multiple sequence alignment.

| Species (Latin) | Species (English) | NCBI Accession number |
|---|---|---|
| **Mammals in conserved site alignment** | | |
| *Homo sapiens* | Human | NP_803875.2 |
| *Pan troglodytes* | Chimpanzee | XP_016815270.4 |
| *Macaca mulatta* | Rhesus macaque | XP_015001362.2 |
| *Lemur catta* | Ring-tailed lemur | XP_045416975.1 |
| *Mus musculus* | Mouse | NP_619615.2 |
| *Rattus norvegicus* | Rat | XP_038935089.1 |
| *Bos taurus* | Cattle | XP_059730442.1 |
| *Canis lupus familiaris* | Dog | XP_038540795.1 |
| **Vertebrates in ConSurf analysis** | | |
| *Acomys russatus* | Golden spiny mouse | XP_051015502.1 |
| *Alexandromys fortis* | Reed vole | XP_050017707.1 |
| *Ambystoma mexicanum* | Axolotl | XP_069478234.1 |
| *Arvicola amphibius* | European water vole | XP_041911647.1 |
| *Balaenoptera acutorostrata* | Common minke whale | XP_057388355.1 |
| *Balaenoptera physalus* | Fin whale | KAB0398262.1 |
| *Bombina bombina* | European fire-bellied toad | XP_053571903.1 |
| *Bos indicus* | Zebu | XP_070658889.1 |
| *Camelus ferus* | Wild Bactrian camel | XP_032323543.1 |
| *Canis lupus familiaris* | Dog | XP_038411278.1 |
| *Carlito syrichta* | Philippine tarsier | XP_021563637.1 |
| *Castor canadensis* | North American beaver | XP_073925166.1 |
| *Cavia porcellus* | Guinea pig | XP_063113705.1 |
| *Chinchilla lanigera* | Long-tailed chinchilla | XP_013368200.1 |
| *Chlorocebus sabaeus* | Green monkey | XP_007999559.3 |
| *Choloepus didactylus* | Linnaeus's two-toed sloth | XP_037658532.1 |
| *Chrysochloris asiatica* | Cape golden mole | XP_006830888.1 |
| *Cricetulus griseus* | Chinese hamster | XP_035310934.1 |
| *Cynocephalus volans* | Philippine flying lemur | XP_062935338.1 |
| *Dasypus novemcinctus* | Nine-banded armadillo | XP_058131902.1 |
| *Diceros bicornis minor* | South-central black rhinoceros | XP_058420460.1 |
| *Dipodomys spectabilis* | Banner-tailed kangaroo rat | XP_042528870.1 |
| *Echinops telfairi* | Lesser hedgehog tenrec | XP_045143035.1 |

| | | |
|---|---|---|
| *Elephantulus edwardii* | Cape elephant shrew | XP_006892412.1 |
| *Elephas maximus indicus* | Indian elephant | XP_049710535.1 |
| *Engystomops pustulosus* | Túngara frog | XP_072007418.1 |
| *Equus asinus* | Donkey | XP_070337588.1 |
| *Erinaceus europaeus* | European hedgehog | XP_060058085.1 |
| *Eulemur rufifrons* | Red-fronted lemur | XP_069321700.1 |
| *Fukomys damarensis* | Damaraland mole-rat | XP_010617839.1 |
| *Galemys pyrenaicus* | Pyrenean desman | KAG8524086.1 |
| *Galeopterus variegatus* | Sunda flying lemur | XP_008588962.1 |
| *Gracilinanus agilis* | Agile gracile opossum | XP_044514196.1 |
| *Gulo gulo* | Wolverine | VCX37434.1 |
| *Heterocephalus glaber* | Naked mole-rat | XP_021118282.1 |
| *Hipposideros armiger* | Great roundleaf bat | XP_019484008.1 |
| *Homo sapiens* | Human | NP_803875.2 |
| *Jaculus jaculus* | Lesser Egyptian jerboa | XP_045000409.1 |
| *Latimeria chalumnae* | Coelacanth | XP_064410285.1 |
| *Lepisosteus oculatus* | Spotted gar | XP_069051252.1 |
| *Macrotis lagotis* | Greater bilby | XP_074057304.1 |
| *Manis pentadactyla* | Chinese pangolin | XP_036732330.2 |
| *Meriones unguiculatus* | Mongolian gerbil | XP_060245461.1 |
| *Miniopterus natalensis* | Natal long-fingered bat | XP_016079739.1 |
| *Mirounga leonina* | Southern elephant seal | KAF3822699.1 |
| *Mixophyes fleayi* | Fleay’s barred frog | XP_075070531.1 |
| *Molossus molossus* | Velvety free-trailed bat | XP_036120365.1 |
| *Monodon monoceros* | Narwhal | TKC41541.1 |
| *Mus musculus* | House mouse | NP_619615.2 |
| *Myodes glareolus* | Bank vole | XP_048313022.1 |
| *Myotis davidii* | David’s myotis | XP_015421619.1 |
| *Myotis myotis* | Greater mouse-eared bat | XP_036193984.1 |
| *Nannospalax galili* | Middle East blind mole-rat | XP_017658290.1 |
| *Ochotona princeps* | American pika | XP_058524528.1 |
| *Octodon degus* | Common degu | XP_004640150.1 |
| *Ornithorhynchus anatinus* | Platypus | XP_028919652.1 |
| *Orycteropus afer afer* | Aardvark | XP_042639560.1 |
| *Oryctolagus cuniculus* | European rabbit | XP_051699933.2 |
| *Otolemur garnettii* | Northern greater galago | XP_023373972.1 |
| *Ovis aries* | Sheep | KAG5204993.1 |
| *Perognathus longimembris pacificus* | Pacific pocket mouse | XP_048215228.1 |
| *Peromyscus maniculatus bairdii* | Eastern deer mouse | XP_015852895.1 |
| *Petaurus breviceps papuanus* | Sugar glider | XP_068937464.1 |
| *Phodopus roborovskii* | Roborovski dwarf hamster | XP_051058465.1 |
| *Phyllostomus discolor* | Pale spear-nosed bat | KAF6100609.1 |
| *Pipistrellus kuhlii* | Kuhl’s pipistrelle | XP_036292686.1 |
| *Plecturocebus cupreus* | Coppery titi monkey | KAL0598757.1 |
| *Pleurodeles waltl* | Iberian ribbed newt | XP_069076731.1 |
| *Pseudophryne corroboree* | Southern corroboree frog | XP_063781339.1 |
| *Pteropus vampyrus* | Large flying fox | XP_039709652.1 |
| *Pyxicephalus adspersus* | African bullfrog | XP_072266834.1 |
| *Rattus rattus* | Black rat | XP_032744443.1 |
| *Rhinolophus ferrumequinum* | Greater horseshoe bat | KAF6323598.1 |
| *Rousettus aegyptiacus* | Egyptian fruit bat | KAF6405462.1 |
| *Saccopteryx leptura* | Lesser sac-winged bat | XP_066235459.1 |
| *Saimiri boliviensis* | Black-capped squirrel monkey | XP_074243158.1 |
| *Sarcophilus harrisii* | Tasmanian devil | XP_031802985.1 |
| *Sciurus carolinensis* | Eastern gray squirrel | XP_047379659.1 |
| *Sigmodon hispidus* | Hispid cotton rat | KAL1767893.1 |
| *Sorex araneus* | Common shrew | XP_054983665.1 |
| *Spea bombifrons* | Plains spadefoot toad | XP_053322520.1 |
| *Suncus etruscus* | Etruscan shrew | XP_049628899.1 |

| | | |
|---|---|---|
| *Suricata suricatta* | Meerkat | XP_029779827.1 |
| *Sus scrofa* | Wild boar | XP_020944671.1 |
| *Talpa occidentalis* | Iberian mole | XP_037358565.1 |
| *Ursus americanus* | American black bear | XP_045661733.1 |
| *Ursus arctos* | Brown bear | XP_048068494.2 |

**Table S3**. **Medications included in the medical treatment for infertility group in the FinnGen analysis.** The treatments can have been used either alone or in variable combinations, hence the combined share of cases exceeds 100%. ATC, anatomical therapeutic chemical classification; FSH, follicle-stimulating hormone.

| ATC code | Name | Description | Share of cases (%) |
|---|---|---|---|
| G03GA01 | Chorionic gonadotrophin | Urine-derived human chorionic gonadotrophin; Ovulation stimulant | 67.93 |
| G03GA02 | Menopausal gonadotrophin | Urine-derived human menopausal gonadotrophin; Ovulation stimulant | 39.67 |
| G03GA04 | Urofollitropin | Urine-derived FSH; Ovulation stimulant | 9.81 |
| G03GA05 | Follitropin alfa | Recombinant FSH; Ovulation stimulant | 46.52 |
| G03GA06 | Follitropin beta | Recombinant FSH; Ovulation stimulant | 38.36 |
| G03GA09 | Corifollitropin alfa | Recombinant FSH; Ovulation stimulant | 4.85 |
| G03GA30 | Combinations | Comprised of combinations of ovulation-stimulative recombinant hormones | 4.73 |
| G03GB02 | Clomifene | Small molecule ovulation stimulant | 19.48 |

**Table S4**. **Analysis of *PKHD1L1* rs17368310 mRNA splicing pattern.** Outcomes of algorithm tools predicting splicing events due to the rs17368310 splice donor variant in intron 47.

| Prediction tool | Event rank | Donor loss | Donor gain | Intronic insertion | Exon skipping | Frameshift |
|---|---|---|---|---|---|---|
| SpliceVault | 1. | - | +22[a], 6.20%[b,c] | CTAATTTTAGCAGCTCTCGTG | - | No |
| | 2. | - | +1556[a], 0.61%[b] | CTAATTTTAGCA…GGTTGTAAAAAA | - | Yes |
| | 3. | - | - | - | Exons 47–50, 0.20%[b] | Yes |
| | 4. | - | - | - | Exons 47–48, 0.06%[b] | Yes |
| | 5. | - | +26[a], 0.03%[b] | CTAATTTTAGCAGCTCTCGTGGTTG | - | Yes |
| | 6. | - | - | - | Exon 47, 0.02%[b] | Yes |
| | 7. | - | - | - | Exons 47–49, 0.02%[b] | Yes |
| | 8. | - | - | - | Exons 46–47, 0.02%[b] | No |
| SpliceRover | 1. | - | +22[a], 0.5287[d] | CTAATTTTAGCAGCTCTCGTG | - | No |
| | 2. | - | +1556[a], 0.4869[d] | CTAATTTTAGCA…GGTTGTAAAAAA | - | Yes |
| SpliceAI | 1. | -1 bp[e], 1.00[f] | 20 bp[e], 0.78[f] | CTAATTTTAGCAGCTCTCGTG | - | No |
| Pangolin | 1. | -1 bp[e], 0.85[f] | 20 bp[e], 0.73[f] | CTAATTTTAGCAGCTCTCGTG | - | No |

[a]Cryptic donor position when the first intronic nucleotide is counted as +1
[b]Prevalence established using RNA-seq samples from Genotype-Tissue Expression and Sequence Read Archive datasets
[c]Whole blood and skeletal muscle
[d]Score describing the relative strength of the donor site
[e]Donor gain position when the SNP is counted as 0
[f]Scores range from 0 to 1 and can be interpreted as the probability that the variant affects splicing at any position within a +/- 500 bp window

**Table S5. Analysis of post-translational modifications in rs17368310-impacted PKHD1L1 protein.** Prediction of post-translational modifications (PTM) on the amino acid insertion ANFSSSR (novel residues inserted between Thr2415–Gly2416) in human PKHD1L1 protein caused by the rs17368310 variant. Prediction confidence threshold, if available, is presented for all theoretically possible PTM events, and score interpretation, *i.e.*, how likely the PTM is to occur, is given. The affected residue is shown in bold, the consensus sequence for the PTM to occur is underlined, and the residues flanking the insertion are shown in italics. ELM, The Eukaryotic Linear Motif resource; GSK3, Glycogen synthase kinase 3.

| PTM | Prediction tool | Residue | Prediction score | Interpretation |
|---|---|---|---|---|
| Methylation | MusiteDeep[a] | …ANFSSS**R***GE*… | 0.362 | Not likely |
| N-glycosylation | ELM[a] | …A**N**FSSSR… | - | Potential |
| | MusiteDeep[a] | …A**N**FSSSR… | 0.908 | Likely |
| | NetNGlyc 1.0[b] | …A**N**FSSSR… | 0.471 (5/9 jury agreement) | Not likely |
| O-glycosylation | MusiteDeep[a] | …ANF**S**SSR… | 0.068 | Not likely |
| | | …ANFS**S**SR… | 0.068 | Not likely |
| | | …ANFSS**S**R… | 0.081 | Not likely |
| | NetOGlyc 4.0[a] | …ANF**S**SSR… | 0.412 | Not likely |
| | | …ANFS**S**SR… | 0.691 | Likely |
| | | …ANFSS**S**R… | 0.307 | Not likely |
| Phosphorylation | ELM[a] | …*D**T***ANFSSSR…[c] | - | Potential (GSK3) |
| | | …ANFSS**S**R*GE*… | - | Potential (Casein kinase 2) |
| | MusiteDeep[a] | …ANF**S**SSR… | 0.303 | Not likely |
| | | …ANFS**S**SR… | 0.268 | Not likely |
| | | …ANFSS**S**R… | 0.412 | Not likely |
| | NetPhos 3.1[b] | …*D**T***ANFSSSR…[c] | 0.458 | Not likely (GSK3) |
| | | …ANF**S**SSR… | 0.969 | Likely (unspecific kinase) |
| | | …ANFS**S**SR… | 0.651 | Likely (unspecific kinase) |
| | | …ANFSS**S**R… | 0.983 | Likely (unspecific kinase) |

[a]The whole variant PKHD1L1 amino acid sequence (4250 amino acids) was used in analysis
[b]The first 4000 amino acids of the variant PKHD1L1 were used in analysis due to program limitations
[c]Novel PTM potentially introduced in the flanking canonical PKHD1L1 protein sequence due to the insertion

**Table S6. *PKHD1L1* locus genetic variants associated with medical treatment for female infertility at genome-wide significance under the recessive model.** Analyses are based on data from the FinnGen project.

| **SNP**[a] | **Position**[b] | **Variant effect prediction** | | | | **Reference/ alternative allele** | **Frequency**[e] | **Additive model genetic association analysis results** | | | **Recessive model genetic association analysis results** | | |
|---|---|---|---|---|---|---|---|---|---|---|---|---|---|
| | | **Consequence** | **Gene** | **IMPACT**[c] | **LOFTEE**[d] | | | **Beta**[e] | **Standard error** | ***p*-value** | **Beta**[e] | **Standard error** | ***p*-value** |
| rs190946682 | 8:109371089 | intron_variant | *PKHD1L1* | MODIFIER | *NA* | G/A | 0.063 | 0.136 | 0.030 | 5.2 x $10^{-6}$ | 0.772 | 0.134 | 8.9 x $10^{-9}$ |
| rs57065002 | 8:109374906 | intron_variant | *PKHD1L1* | MODIFIER | *NA* | C/T | 0.067 | 0.134 | 0.029 | 4.0 x $10^{-6}$ | 0.752 | 0.126 | 2.5 x $10^{-9}$ |
| rs4735131 | 8:109381753 | intron_variant | *PKHD1L1* | MODIFIER | *NA* | C/T | 0.067 | 0.134 | 0.029 | 3.8 x $10^{-6}$ | 0.754 | 0.126 | 2.3 x $10^{-9}$ |
| rs12164209 | 8:109407609 | intron_variant | *PKHD1L1* | MODIFIER | *NA* | C/G | 0.067 | 0.134 | 0.029 | 4.1 x $10^{-6}$ | 0.753 | 0.126 | 2.5 x $10^{-9}$ |
| rs16879547 | 8:109430154 | intron_variant | *PKHD1L1* | MODIFIER | *NA* | C/G | 0.067 | 0.135 | 0.029 | 3.4 x $10^{-6}$ | 0.751 | 0.127 | 3.3 x $10^{-9}$ |
| rs9643050 | 8:109458129 | intron_variant | *PKHD1L1* | MODIFIER | *NA* | T/C | 0.068 | 0.140 | 0.029 | 1.1 x $10^{-6}$ | 0.730 | 0.124 | 4.2 x $10^{-9}$ |
| **rs17368310** | **8:109459837** | **splice_donor _variant** | ***PKHD1L1*** | **HIGH** | **High confidence** | **G/C** | **0.068** | **0.140** | **0.029** | **1.1 x $10^{-6}$** | **0.729** | **0.124** | **4.3 x $10^{-9}$** |
| rs1964514 | 8:109463457 | intron_variant | *PKHD1L1* | MODIFIER | *NA* | G/C | 0.068 | 0.140 | 0.029 | 1.1 x $10^{-6}$ | 0.730 | 0.124 | 4.2 x $10^{-9}$ |
| rs12549027 | 8:109475283 | intron_variant | *PKHD1L1* | MODIFIER | *NA* | T/A | 0.068 | 0.140 | 0.029 | 1.1 x $10^{-6}$ | 0.730 | 0.124 | 4.2 x $10^{-9}$ |
| chr8:1094931 74:C:CAT | 8:109493174 | *NA (intronic)* | *PKHD1L1* | *NA* | *NA* | C/CAT | 0.068 | 0.140 | 0.029 | 1.1 x $10^{-6}$ | 0.730 | 0.124 | 4.6 x $10^{-9}$ |
| rs58870933 | 8:109515401 | intron_variant | *PKHD1L1* | MODIFIER | *NA* | CAAT/C | 0.068 | 0.140 | 0.029 | 1.1 x $10^{-6}$ | 0.732 | 0.124 | 3.8 x $10^{-9}$ |
| rs147615814 | 8:109563649 | intron_variant | *EBAG9* | MODIFIER | *NA* | CAT/C | 0.071 | 0.133 | 0.028 | 2.7 x $10^{-6}$ | 0.717 | 0.121 | 3.0 x $10^{-9}$ |
| rs17378154 | 8:109568721 | 3_prime_UTR _variant, downstream_ gene_variant | *EBAG9* | MODIFIER | *NA* | G/A | 0.068 | 0.138 | 0.029 | 1.6 x $10^{-6}$ | 0.726 | 0.124 | 5.2 x $10^{-9}$ |

[a]All genome-wide significant SNPs shown
[b]Position of SNP in the human genome (chromosome:position)
[c]VEP (Variant Effect Predictor) impact MODIFIER refers to non-coding variants or variants affecting non-coding genes, where predictions are difficult or there is no evidence of impact; HIGH refers to a variant assumed to have high (disruptive) impact in the protein, probably leading to protein truncation, loss-of-function or nonsense-mediated decay
[d]LOFTEE (Loss-Of-Function Transcript Effect Estimator) prediction of being a loss-of-function variant
[e]Frequency and beta shown for the alternative allele

**Table S7. Having children according to *PKHD1L1* rs17368310 genotypes.** FinnGen women who had reached the age of 45 years were included. Information on the number of children was extracted from the Finnish Population and Medical Birth Registries.

| Group | All genotypes | | CC homozygotes | | CG heterozygotes | | GG homozygotes | | *p*-value[a], CC vs other genotypes |
|---|---|---|---|---|---|---|---|---|---|
| | Total *n* | Does not have children, *n* (%) | Total *n* | Does not have children, *n* (%) | Total *n* | Does not have children, *n* (%) | Total *n* | Does not have children, *n* (%) | |
| All women | 186631 | 28766 (15.4) | 897 | 170 (19.0) | 23467 | 3738 (15.9) | 162267 | 23763 (15.3) | $3.78 \times 10^{-3}$ |
| Women with medical treatment for infertility | 5522 | 1291 (23.4) | 49 | 22 (44.9) | 746 | 174 (23.3) | 4727 | 1095 (23.2) | $6.61 \times 10^{-4}$ |
| Control women | 181109 | 27475 (15.2) | 848 | 148 (17.5) | 22721 | 3564 (15.7) | 157540 | 23763 (15.1) | 0.070 |

[a]χ2 test

**Table S8. Number of children according to *PKHD1L1* rs17368310 genotypes.** FinnGen women who had reached the age of 45 years were included. Information on the number of children was extracted from the Finnish Population and Medical Birth Registries.

| Group | All genotypes | | CC homozygotes | | CG heterozygotes | | GG homozygotes | | *p*-value[a], CC vs other genotypes |
|---|---|---|---|---|---|---|---|---|---|
| | Total *n* | Mean number of children (95%CI) | Total *n* | Mean number of children (95%CI) | Total *n* | Mean number of children (95%CI) | Total *n* | Mean number of children (95%CI) | |
| All women | 186631 | 1.94 (1.93-1.95) | 897 | 1.78 (1.70-1.87) | 23467 | 1.93 (1.91-1.95) | 162267 | 1.94 (1.94-1.95) | $3.11 \times 10^{-3}$ |
| Women with medical treatment for infertility | 5522 | 1.52 (1.49-1.55) | 49 | 0.94 (0.65-1.23) | 746 | 1.54 (1.45-1.63) | 4727 | 1.52 (1.49-1.56) | $1.78 \times 10^{-4}$ |
| Control women | 181109 | 1.95 (1.95-9.96) | 848 | 1.83 (1.75-1.92) | 22721 | 1.94 (1.92-1.96) | 157540 | 1.96 (1.95-1.96) | $6.97 \times 10^{-3}$ |

[a]Independent samples *t*-test

**Table S9. Number of pregnancies according to *PKHD1L1* rs17368310 genotypes.** Data from the Mexico City Prospective Study was used. Information on the number of pregnancies is based on study questionnaires.

| Group/analysis | CC homozygotes | | CG heterozygotes | | GG homozygotes | | *p*-value[a], CC vs other genotypes | Genetic association analysis, additive model results | | | Genetic association analysis, recessive model results | | |
|---|---|---|---|---|---|---|---|---|---|---|---|---|---|
| | Total *n* | Mean number of pregnancies (95%CI) | Total *n* | Mean number of pregnancies (95%CI) | Total *n* | Mean number of pregnancies (95%CI) | | Beta[b] | Standard error | *p*-value | Beta[b] | Standard error | *p*-value |
| Mexico City Prospective Study women | 9637 | 4.73 (4.66-4.80) | 39547 | 4.85 (4.82-4.88) | 44929 | 4.80 (4.77-4.83) | $4.4 \times 10^{-3}$ | -0.0258 | 0.0045 | $7.3 \times 10^{-9}$ | -0.070 | 0.0097 | $6.1 \times 10^{-13}$ |

[a]Independent samples *t*-test
[b]Betas with reference to rs17368310 C allele; Frequency of C allele was 0.311

**Table S10.** FinnGen project author list.

| FinnGen | | | |
|---|---|---|---|
| **Full Name** | **Affiliation** | **Role 1** | **Role 2** |
| Aarno Palotie | Institute for Molecular Medicine Finland (FIMM), HiLIFE, University of Helsinki, Helsinki, Finland; Broad Institute of MIT and Harvard; Massachusetts General Hospital | Steering Committee | Steering Committee |
| Mark Daly | Institute for Molecular Medicine Finland (FIMM), HiLIFE, University of Helsinki, Helsinki, Finland; Broad Institute of MIT and Harvard; Massachusetts General Hospital | Steering Committee | Steering Committee |
| Bridget Riley-Gillis | Abbvie, Chicago, IL, United States | Steering Committee | Pharmaceutical companies |
| Howard Jacob | Abbvie, Chicago, IL, United States | Steering Committee | Pharmaceutical companies |
| Coralie Viollet | AstraZeneca, Cambridge, United Kingdom | Steering Committee | Pharmaceutical companies |
| Slavé Petrovski | AstraZeneca, Cambridge, United Kingdom | Steering Committee | Pharmaceutical companies |
| Alix Berton | Bayer AG, Leverkusen, Germany | Steering Committee | Pharmaceutical companies |
| Santha Ramakrishnan | Bayer AG, Leverkusen, Germany | Steering Committee | Pharmaceutical companies |
| Ellen Tsai | Biogen, Cambridge, MA, United States | Steering Committee | Pharmaceutical companies |
| Zhihao Ding | Boehringer Ingelheim, Ingelheim am Rhein, Germany | Steering Committee | Pharmaceutical companies |
| Emily Holzinger | Bristol Myers Squibb, New York, NY, United States | Steering Committee | Pharmaceutical companies |
| Robert Plenge | Bristol Myers Squibb, New York, NY, United States | Steering Committee | Pharmaceutical companies |
| Joseph Maranville | Bristol Myers Squibb, New York, NY, United States | Steering Committee | Pharmaceutical companies |
| Mark McCarthy | Genentech, San Francisco, CA, United States | Steering Committee | Pharmaceutical companies |
| Rion Pendergrass | Genentech, San Francisco, CA, United States | Steering Committee | Pharmaceutical companies |
| Jonathan Davitte | GlaxoSmithKline, Collegeville, PA, United States | Steering Committee | Pharmaceutical companies |
| Chia-Yen Chen | Merck, Kenilworth, NJ, United States | Steering Committee | Pharmaceutical companies |
| Melis Atalar Aksit | Pfizer, New York, NY, United States | Steering Committee | Pharmaceutical companies |
| Anna Vlahiotis | Pfizer, New York, NY, United States | Steering Committee | Pharmaceutical companies |
| Katherine Klinger | Translational Sciences, Sanofi R&D, Framingham, MA, USA | Steering Committee | Pharmaceutical companies |
| Clement Chatelain | Translational Sciences, Sanofi R&D, Framingham, MA, USA | Steering Committee | Pharmaceutical companies |
| Jorg Blankenstein | Translational Sciences, Sanofi R&D, Framingham, MA, USA | Steering Committee | Pharmaceutical companies |
| Karol Estrada | Maze Therapeutics, San Francisco, CA, United States | Steering Committee | Pharmaceutical companies |
| Robert Graham | Maze Therapeutics, San Francisco, CA, United States | Steering Committee | Pharmaceutical companies |
| Dawn Waterworth | Johnson & Johnson Innovative Medicine, Spring House, PA, United States | Steering Committee | Pharmaceutical companies |
| Chris O´Donnell | Novartis Institutes for BioMedical Research, Cambridge, MA, United States | Steering Committee | Pharmaceutical companies |
| Nicole Renaud | Novartis Institutes for BioMedical Research, Cambridge, MA, United States | Steering Committee | Pharmaceutical companies |
| Tomi P. Mäkelä | HiLIFE, University of Helsinki, Finland, Finland | Steering Committee | University of Helsinki & Biobanks |
| Jaakko Kaprio | Institute for Molecular Medicine Finland (FIMM), HiLIFE, University of Helsinki, Helsinki, Finland | Steering Committee | University of Helsinki & Biobanks |
| Minna Ruddock | Arctic biobank / University of Oulu | Steering Committee | University of Helsinki & Biobanks |
| Lila Kallio | Auria Biobank / University of Turku / Wellbeing Services County of Southwest Finland, Turku, Finland | Steering Committee | University of Helsinki & Biobanks |
| Antti Hakanen | Auria Biobank / University of Turku / Wellbeing Services County of Southwest Finland, Turku, Finland | Steering Committee | University of Helsinki & Biobanks |
| Terhi Kilpi | THL Biobank / Finnish Institute for Health and Welfare (THL), Helsinki, Finland | Steering Committee | University of Helsinki & Biobanks |
| Markus Perola | THL Biobank / Finnish Institute for Health and Welfare (THL), Helsinki, Finland | Steering Committee | University of Helsinki & Biobanks |
| Jukka Partanen | Finnish Red Cross Blood Service / Finnish Hematology Registry and Clinical Biobank, Helsinki, Finland | Steering Committee | University of Helsinki & Biobanks |
| Taneli Raivio | Helsinki Biobank / Helsinki University and Hospital District of Helsinki and Uusimaa, Helsinki | Steering Committee | University of Helsinki & Biobanks |
| Eero Punkka | Helsinki Biobank / Helsinki University and Hospital District of Helsinki and Uusimaa, Helsinki | Steering Committee | University of Helsinki & Biobanks |
| Teija Kekonen | Northern Finland Biobank Borealis / University of Oulu / Wellbeing services county of North Ostrobothnia, Oulu, Finland | Steering Committee | University of Helsinki & Biobanks |
| Raisa Serpi | Northern Finland Biobank Borealis / University of Oulu / Wellbeing services county of North Ostrobothnia, Oulu, Finland | Steering Committee | University of Helsinki & Biobanks |
| Kati Kristiansson | Finnish Clinical Biobank Tampere / University of Tampere / Wellbeing Services County of Pirkanmaa, Tampere, Finland | Steering Committee | University of Helsinki & Biobanks |
| Sanna Siltanen | Finnish Clinical Biobank Tampere / University of Tampere / Wellbeing Services County of Pirkanmaa, Tampere, Finland | Steering Committee | University of Helsinki & Biobanks |
| Veli-Matti Kosma | Biobank of Eastern Finland / University of Eastern Finland / Wellbeing services county of North Savo, Kuopio, Finland | Steering Committee | University of Helsinki & Biobanks |
| Arto Mannermaa | Biobank of Eastern Finland / University of Eastern Finland / Wellbeing services county of North Savo, Kuopio, Finland | Steering Committee | University of Helsinki & Biobanks |
| Jari Laukkanen | Central Finland Biobank / University of Jyväskylä / Wellbeing Services County of Central Finland, Jyväskylä, Finland | Steering Committee | University of Helsinki & Biobanks |
| Tiina Jokela | Central Finland Biobank / University of Jyväskylä / Wellbeing Services County of Central Finland, Jyväskylä, Finland | Steering Committee | University of Helsinki & Biobanks |
| Mervi Ahlroth | Finnish Biobank Cooperative - FINBB | Steering Committee | University of Helsinki & Biobanks |
| Johanna Mäkelä | Finnish Biobank Cooperative - FINBB | Steering Committee | University of Helsinki & Biobanks |
| Outi Tuovila | Business Finland, Helsinki, Finland | Steering Committee | Other Experts/ Non-Voting Members |
| Jeffrey Waring | Abbvie, Chicago, IL, United States | Scientific Committee | Pharmaceutical companies |
| Bridget Riley-Gillis | Abbvie, Chicago, IL, United States | Scientific Committee | Pharmaceutical companies |
| Fedik Rahimov | Abbvie, Chicago, IL, United States | Scientific Committee | Pharmaceutical companies |
| Ioanna Tachmazidou | AstraZeneca, Cambridge, United Kingdom | Scientific Committee | Pharmaceutical companies |
| Slavé Petrovski | AstraZeneca, Cambridge, United Kingdom | Scientific Committee | Pharmaceutical companies |
| Alix Berton | Bayer AG, Leverkusen, Germany | Scientific Committee | Pharmaceutical companies |
| Santha Ramakrishnan | Bayer AG, Leverkusen, Germany | Scientific Committee | Pharmaceutical companies |
| Ellen Tsai | Biogen, Cambridge, MA, United States | Scientific Committee | Pharmaceutical companies |
| Zhihao Ding | Boehringer Ingelheim, Ingelheim am Rhein, Germany | Scientific Committee | Pharmaceutical companies |
| Marc Jung | Boehringer Ingelheim, Ingelheim am Rhein, Germany | Scientific Committee | Pharmaceutical companies |
| Hanati Tuoken | Boehringer Ingelheim, Ingelheim am Rhein, Germany | Scientific Committee | Pharmaceutical companies |
| Shameek Biswas | Bristol Myers Squibb, New York, NY, United States | Scientific Committee | Pharmaceutical companies |
| Benjamin Sun | Bristol Myers Squibb, New York, NY, United States | Scientific Committee | Pharmaceutical companies |
| Rion Pendergrass | Genentech, San Francisco, CA, United States | Scientific Committee | Pharmaceutical companies |
| Jonathan Davitte | GlaxoSmithKline, Collegeville, PA, United States | Scientific Committee | Pharmaceutical companies |
| Neha Raghavan | Merck, Kenilworth, NJ, United States | Scientific Committee | Pharmaceutical companies |
| Jae-Hoon Sul | Merck, Kenilworth, NJ, United States | Scientific Committee | Pharmaceutical companies |
| Melis Atalar Aksit | Pfizer, New York, NY, United States | Scientific Committee | Pharmaceutical companies |
| Xinli Hu | Pfizer, New York, NY, United States | Scientific Committee | Pharmaceutical companies |
| Katherine Klinger | Translational Sciences, Sanofi R&D, Framingham, MA, USA | Scientific Committee | Pharmaceutical companies |
| Robert Graham | Maze Therapeutics, San Francisco, CA, United States | Scientific Committee | Pharmaceutical companies |
| Dawn Waterworth | Johnson & Johnson Innovative Medicine, Spring House, PA, United States | Scientific Committee | Pharmaceutical companies |
| Nicole Renaud | Novartis Institutes for BioMedical Research, Cambridge, MA, United States | Scientific Committee | Pharmaceutical companies |
| Ma´en Obeidat | Novartis Institutes for BioMedical Research, Cambridge, MA, United States | Scientific Committee | Pharmaceutical companies |
| Jonathan Chung | Novartis Institutes for BioMedical Research, Cambridge, MA, United States | Scientific Committee | Pharmaceutical companies |
| Jonas Zierer | Novartis Institutes for BioMedical Research, Cambridge, MA, United States | Scientific Committee | Pharmaceutical companies |
| Mari Niemi | Novartis Institutes for BioMedical Research, Cambridge, MA, United States | Scientific Committee | Pharmaceutical companies |
| Samuli Ripatti | Institute for Molecular Medicine Finland (FIMM), HiLIFE, University of Helsinki, Helsinki, Finland | Scientific Committee | University of Helsinki & Biobanks |
| Johanna Schleutker | Auria Biobank / University of Turku / Wellbeing Services County of Southwest Finland, Turku, Finland | Scientific Committee | University of Helsinki & Biobanks |
| Markus Perola | THL Biobank / Finnish Institute for Health and Welfare (THL), Helsinki, Finland | Scientific Committee | University of Helsinki & Biobanks |
| Tiina Wahlfors | THL Biobank / Finnish Institute for Health and Welfare (THL), Helsinki, Finland | Scientific Committee | University of Helsinki & Biobanks |
| Mikko Arvas | Finnish Red Cross Blood Service / Finnish Hematology Registry and Clinical Biobank, Helsinki, Finland | Scientific Committee | University of Helsinki & Biobanks |
| Olli Carpén | Helsinki Biobank / Helsinki University and Hospital District of Helsinki and Uusimaa, Helsinki | Scientific Committee | University of Helsinki & Biobanks |
| Reetta Hinttala | Northern Finland Biobank Borealis / University of Oulu / Wellbeing services county of North Ostrobothnia, Oulu, Finland | Scientific Committee | University of Helsinki & Biobanks |

| | | | |
|---|---|---|---|
| Johannes Kettunen | Northern Finland Biobank Borealis / University of Oulu / Wellbeing services county of North Ostrobothnia, Oulu, Finland | Scientific Committee | University of Helsinki & Biobanks |
| Arto Mannermaa | Biobank of Eastern Finland / University of Eastern Finland / Wellbeing services county of North Savo, Kuopio, Finland | Scientific Committee | University of Helsinki & Biobanks |
| Katriina Aalto-Setälä | Faculty of Medicine and Health Technology, Tampere University, Tampere, Finland | Scientific Committee | University of Helsinki & Biobanks |
| Mika Kähönen | Finnish Clinical Biobank Tampere / University of Tampere / Wellbeing Services County of Pirkanmaa, Tampere, Finland | Scientific Committee | University of Helsinki & Biobanks |
| Jari Laukkanen | Central Finland Biobank / University of Jyväskylä / Wellbeing Services County of Central Finland, Jyväskylä, Finland | Scientific Committee | University of Helsinki & Biobanks |
| Johanna Mäkelä | FINBB - Finnish biobank cooperative | Scientific Committee | University of Helsinki & Biobanks |
| Hanna Kujala | Biobank of Eastern Finland / University of Eastern Finland / Wellbeing services county of North Savo, Kuopio, Finland | Clinical Group / Task Force | |
| Triin Laisk | Estonian biobank, Tartu, Estonia | Clinical Group / Task Force | |
| Natalia Pujol | Estonian biobank, Tartu, Estonia | Clinical Group / Task Force | |
| Mika Kähönen | Finnish Clinical Biobank Tampere / University of Tampere / Wellbeing Services County of Pirkanmaa, Tampere, Finland | Clinical Group / Task Force | |
| Veikko Salomaa | Finnish Institute for Health and Welfare (THL), Helsinki, Finland | Clinical Group / Task Force | |
| Jaana Suvisaari | Finnish Institute for Health and Welfare (THL), Helsinki, Finland | Clinical Group / Task Force | |
| Satu Koskela | Finnish Red Cross Blood Service / Finnish Hematology Registry and Clinical Biobank, Helsinki, Finland | Clinical Group / Task Force | |
| Jouni Lauronen | Finnish Red Cross Blood Service / Finnish Hematology Registry and Clinical Biobank, Helsinki, Finland | Clinical Group / Task Force | |
| Kristiina Aittomäki | Helsinki University Central Hospital, Helsinki, Finland | Clinical Group / Task Force | |
| Pirkko Pussinen | Helsinki University Hospital and University of Helsinki, Helsinki / University of Eastern Finland. Kuopio, Finland | Clinical Group / Task Force | |
| Tuomo Meretoja | Helsinki University Hospital and University of Helsinki, Helsinki, Finland | Clinical Group / Task Force | |
| Heikki Joensuu | Helsinki University Hospital and University of Helsinki, Helsinki, Finland | Clinical Group / Task Force | |
| Peeter Karihtala | Helsinki University Hospital and University of Helsinki, Helsinki, Finland | Clinical Group / Task Force | |
| Emma Juuri | Helsinki University Hospital and University of Helsinki, Helsinki, Finland | Clinical Group / Task Force | |
| Aino Salminen | Helsinki University Hospital and University of Helsinki, Helsinki, Finland | Clinical Group / Task Force | |
| Tuula Salo | Helsinki University Hospital and University of Helsinki, Helsinki, Finland | Clinical Group / Task Force | |
| David Rice | Helsinki University Hospital and University of Helsinki, Helsinki, Finland | Clinical Group / Task Force | |
| Pekka Nieminen | Helsinki University Hospital and University of Helsinki, Helsinki, Finland | Clinical Group / Task Force | |
| Ulla Palotie | Helsinki University Hospital and University of Helsinki, Helsinki, Finland | Clinical Group / Task Force | |
| Fredrik Åberg | Helsinki University Hospital and University of Helsinki, Helsinki, Finland | Clinical Group / Task Force | |
| Daniel Gordin | Helsinki University Hospital and University of Helsinki, Helsinki, Finland | Clinical Group / Task Force | |
| Patrik Finne | Helsinki University Hospital and University of Helsinki, Helsinki, Finland | Clinical Group / Task Force | |
| Joni A Turunen | Helsinki University Hospital and University of Helsinki, Helsinki, Finland; Folkhälsan Research Center, Helsinki, Finland | Clinical Group / Task Force | |
| Minna Raivio | Hospital District of Helsinki and Uusimaa, Helsinki, Finland | Clinical Group / Task Force | |
| Pentti Tienari | Hospital District of Helsinki and Uusimaa, Helsinki, Finland | Clinical Group / Task Force | |
| Martti Färkkilä | Hospital District of Helsinki and Uusimaa, Helsinki, Finland | Clinical Group / Task Force | |
| Jukka Koskela | Hospital District of Helsinki and Uusimaa, Helsinki, Finland | Clinical Group / Task Force | |
| Sampsa Pikkarainen | Hospital District of Helsinki and Uusimaa, Helsinki, Finland | Clinical Group / Task Force | |
| Kari Eklund | Hospital District of Helsinki and Uusimaa, Helsinki, Finland | Clinical Group / Task Force | |
| Paula Kauppi | Hospital District of Helsinki and Uusimaa, Helsinki, Finland | Clinical Group / Task Force | |
| Daniel Gordin | Hospital District of Helsinki and Uusimaa, Helsinki, Finland | Clinical Group / Task Force | |
| Juha Sinisalo | Hospital District of Helsinki and Uusimaa, Helsinki, Finland | Clinical Group / Task Force | |
| Marja-Riitta Taskinen | Hospital District of Helsinki and Uusimaa, Helsinki, Finland | Clinical Group / Task Force | |
| Tiinamaija Tuomi | Hospital District of Helsinki and Uusimaa, Helsinki, Finland | Clinical Group / Task Force | |
| Timo Hiltunen | Hospital District of Helsinki and Uusimaa, Helsinki, Finland | Clinical Group / Task Force | |
| Johanna Mattson | Hospital District of Helsinki and Uusimaa, Helsinki, Finland | Clinical Group / Task Force | |
| Eveliina Salminen | Hospital District of Helsinki and Uusimaa, Helsinki, Finland | Clinical Group / Task Force | |
| Terhi Ollila | Hospital District of Helsinki and Uusimaa, Helsinki, Finland | Clinical Group / Task Force | |
| Katariina Hannula-Jouppi | Hospital District of Helsinki and Uusimaa, Helsinki, Finland | Clinical Group / Task Force | |
| Oskari Heikinheimo | Hospital District of Helsinki and Uusimaa, Helsinki, Finland | Clinical Group / Task Force | |
| Ilkka Kalliala | Hospital District of Helsinki and Uusimaa, Helsinki, Finland | Clinical Group / Task Force | |
| Lauri Aaltonen | Hospital District of Helsinki and Uusimaa, Helsinki, Finland | Clinical Group / Task Force | |
| Erkki Isometsä | Hospital District of Helsinki and Uusimaa, Helsinki, Finland | Clinical Group / Task Force | |
| Antti Aarnisalo | Hospital District of Helsinki and Uusimaa, Helsinki, Finland | Clinical Group / Task Force | |
| Ilkka Immonen | Hospital District of Helsinki and Uusimaa, Helsinki, Finland | Clinical Group / Task Force | |
| Salla Ranta | Hospital District of Helsinki and Uusimaa, Helsinki, Finland | Clinical Group / Task Force | |
| Filip Scheperjans | Hospital District of Helsinki and Uusimaa, Helsinki, Finland | Clinical Group / Task Force | |
| Felix Vaura | Institute for Molecular Medicine Finland (FIMM), HiLIFE, University of Helsinki, Helsinki, | Clinical Group / Task Force | |
| Nina Mars | Institute for Molecular Medicine Finland (FIMM), HiLIFE, University of Helsinki, Helsinki, | Clinical Group / Task Force | |
| Esa Pitkänen | Institute for Molecular Medicine Finland (FIMM), HiLIFE, University of Helsinki, Helsinki, | Clinical Group / Task Force | |
| Hannele Laivuori | Institute for Molecular Medicine Finland (FIMM), HiLIFE, University of Helsinki, Helsinki, | Clinical Group / Task Force | |
| Tuomo Kiiskinen | Institute for Molecular Medicine Finland (FIMM), HiLIFE, University of Helsinki, Helsinki, Finland | Clinical Group / Task Force | |
| Katja Kivinen | Institute for Molecular Medicine Finland (FIMM), HiLIFE, University of Helsinki, Helsinki, | Clinical Group / Task Force | |
| Elisabeth Widen | Institute for Molecular Medicine Finland (FIMM), HiLIFE, University of Helsinki, Helsinki, | Clinical Group / Task Force | |
| Taru Tukiainen | Institute for Molecular Medicine Finland (FIMM), HiLIFE, University of Helsinki, Helsinki, | Clinical Group / Task Force | |
| Hanna Ollila | Institute for Molecular Medicine Finland (FIMM), HiLIFE, University of Helsinki, Helsinki, | Clinical Group / Task Force | |
| Elmo Saarentaus | Institute for Molecular Medicine Finland (FIMM), HiLIFE, University of Helsinki, Helsinki, | Clinical Group / Task Force | |
| Anne Kerola | Institute for Molecular Medicine Finland (FIMM), HiLIFE, University of Helsinki, Helsinki, | Clinical Group / Task Force | |
| Eero Vuoksimaa | Institute for Molecular Medicine Finland (FIMM), HiLIFE, University of Helsinki, Helsinki, | Clinical Group / Task Force | |
| Joni Lindbohm | Institute for Molecular Medicine Finland (FIMM), HiLIFE, University of Helsinki, Helsinki, | Clinical Group / Task Force | |
| Zhiyu Yang | Institute for Molecular Medicine Finland (FIMM), HiLIFE, University of Helsinki, Helsinki, | Clinical Group / Task Force | |
| Matthew Sampson | Institute for Molecular Medicine Finland (FIMM), HiLIFE, University of Helsinki, Helsinki, Finland; Broad Institute & Harvard Medical School, Cambridge, United States | Clinical Group / Task Force | |
| Michelle McNulty | Institute for Molecular Medicine Finland (FIMM), HiLIFE, University of Helsinki, Helsinki, Finland; Broad Institute & Harvard Medical School, Cambridge, United States | Clinical Group / Task Force | |
| Aoxing Liu | Institute for Molecular Medicine Finland (FIMM), HiLIFE, University of Helsinki, Helsinki, Finland; Broad Institute, Cambridge, MA, United States | Clinical Group / Task Force | |
| Joel Rämö | Institute for Molecular Medicine Finland (FIMM), HiLIFE, University of Helsinki, Helsinki, Finland; Broad Institute, Cambridge, MA, United States | Clinical Group / Task Force | |
| Austin Argentieri | Institute for Molecular Medicine Finland (FIMM), HiLIFE, University of Helsinki, Helsinki, Finland; Broad Institute, Cambridge, MA, United States | Clinical Group / Task Force | |
| Amanda Elliott | Institute for Molecular Medicine Finland (FIMM), HiLIFE, University of Helsinki, Helsinki, Finland; Broad Institute, Cambridge, MA, USA and Massachusetts General Hospital, Boston, MA, USA | Clinical Group / Task Force | |
| Elisa Rahikkala | Northern Ostrobothnia Hospital District, Oulu, Finland | Clinical Group / Task Force | |
| Kirsi Sipilä | Oulu University Hospital and University of Oulu, Oulu, Finland | Clinical Group / Task Force | |
| Valtteri Julkunen | University of Eastern Finland and Kuopio University Hospital, Kuopio, Finland | Clinical Group / Task Force | |
| Ville Leinonen | University of Eastern Finland and Kuopio University Hospital, Kuopio, Finland | Clinical Group / Task Force | |
| Sanna Toppila-Salmi | University of Eastern Finland and Kuopio University Hospital, Kuopio, Finland; Helsinki University Hospital and University of Helsinki, Finland | Clinical Group / Task Force | |
| Mikko Hiltunen | University of Eastern Finland, Kuopio, Finland | Clinical Group / Task Force | |
| Eino Solje | University of Eastern Finland, Kuopio, Finland | Clinical Group / Task Force | |
| Hannu Kankaanranta | University of Gothenburg, Gothenburg, Sweden/ Seinäjoki Central Hospital, Seinäjoki, Finland/ Tampere University, Tampere, Finland | Clinical Group / Task Force | |
| Antti Mäkitie | University of Helsinki and Helsinki University Hospital, Helsinki, Finland | Clinical Group / Task Force | |
| Iiris Hovatta | University of Helsinki, Helsinki, Finland | Clinical Group / Task Force | |
| Niko Välimäki | University of Helsinki, Helsinki, Finland | Clinical Group / Task Force | |
| Minttu Marttila | University of Helsinki, Helsinki, Finland | Clinical Group / Task Force | |

| | | |
|---|---|---|
| Anne Portaankorva | University of Helsinki, Helsinki, Finland | Clinical Group / Task Force |
| Eija Laakkonen | University of Jyväskylä, Jyväskylä, Finland | Clinical Group / Task Force |
| Heidi Silven | University of Oulu, Oulu, Finland | Clinical Group / Task Force |
| Eeva Sliz | University of Oulu, Oulu, Finland | Clinical Group / Task Force |
| Riikka Arffman | University of Oulu, Oulu, Finland | Clinical Group / Task Force |
| Susanna Savukoski | University of Oulu, Oulu, Finland | Clinical Group / Task Force |
| Riitta Kaarteenaho | University of Oulu, Oulu, Finland | Clinical Group / Task Force |
| Jaakko Tyrmi | University of Oulu, Oulu, Finland / University of Tampere, Tampere, Finland | Clinical Group / Task Force |
| Laura Kuusalo | University of Turku, Turku, Finland | Clinical Group / Task Force |
| Laura Pirilä | University of Turku, Turku, Finland | Clinical Group / Task Force |
| Tapio Hellman | University of Turku, Turku, Finland | Clinical Group / Task Force |
| Matti Vuori | University of Turku, Turku, Finland | Clinical Group / Task Force |
| Teemu Niiranen | University of Turku, Turku, Finland; Finnish Institute for Health and Welfare (THL), Helsinki, Finland | Clinical Group / Task Force |
| Timo Blomster | Wellbeing services county of North Ostrobothnia, Oulu, Finland | Clinical Group / Task Force |
| Johanna Huhtakangas | Wellbeing services county of North Ostrobothnia, Oulu, Finland | Clinical Group / Task Force |
| Terttu Harju | Wellbeing services county of North Ostrobothnia, Oulu, Finland | Clinical Group / Task Force |
| Kaisa Tasanen | Wellbeing services county of North Ostrobothnia, Oulu, Finland | Clinical Group / Task Force |
| Laura Huilaja | Wellbeing services county of North Ostrobothnia, Oulu, Finland | Clinical Group / Task Force |
| Vuokko Anttonen | Wellbeing services county of North Ostrobothnia, Oulu, Finland | Clinical Group / Task Force |
| Marja Vääräsmäki | Wellbeing services county of North Ostrobothnia, Oulu, Finland | Clinical Group / Task Force |
| Outi Uimari | Wellbeing services county of North Ostrobothnia, Oulu, Finland | Clinical Group / Task Force |
| Laure Morin-Papunen | Wellbeing services county of North Ostrobothnia, Oulu, Finland | Clinical Group / Task Force |
| Maarit Niinimäki | Wellbeing services county of North Ostrobothnia, Oulu, Finland | Clinical Group / Task Force |
| Terhi Piltonen | Wellbeing services county of North Ostrobothnia, Oulu, Finland | Clinical Group / Task Force |
| Reetta Kälviäinen | Wellbeing services county of North Savo, Kuopio, Finland | Clinical Group / Task Force |
| Valtteri Julkunen | Wellbeing services county of North Savo, Kuopio, Finland | Clinical Group / Task Force |
| Hilkka Soininen | Wellbeing services county of North Savo, Kuopio, Finland | Clinical Group / Task Force |
| Mikko Kiviniemi | Wellbeing services county of North Savo, Kuopio, Finland | Clinical Group / Task Force |
| Oili Kaipiainen-Seppänen | Wellbeing services county of North Savo, Kuopio, Finland | Clinical Group / Task Force |
| Margit Pelkonen | Wellbeing services county of North Savo, Kuopio, Finland | Clinical Group / Task Force |
| Päivi Auvinen | Wellbeing services county of North Savo, Kuopio, Finland | Clinical Group / Task Force |
| Maria Siponen | Wellbeing services county of North Savo, Kuopio, Finland | Clinical Group / Task Force |
| Liisa Suominen | Wellbeing services county of North Savo, Kuopio, Finland | Clinical Group / Task Force |
| Päivi Mäntylä | Wellbeing services county of North Savo, Kuopio, Finland | Clinical Group / Task Force |
| Kai Kaarniranta | Wellbeing services county of North Savo, Kuopio, Finland; University of Lodz, Lodz, Poland | Clinical Group / Task Force |
| Jukka Peltola | Wellbeing Services County of Pirkanmaa, Tampere, Finland | Clinical Group / Task Force |
| Airi Jussila | Wellbeing Services County of Pirkanmaa, Tampere, Finland | Clinical Group / Task Force |
| Katri Kaukinen | Wellbeing Services County of Pirkanmaa, Tampere, Finland | Clinical Group / Task Force |
| Pia Isomäki | Wellbeing Services County of Pirkanmaa, Tampere, Finland | Clinical Group / Task Force |
| Jussi Hernesniemi | Wellbeing Services County of Pirkanmaa, Tampere, Finland | Clinical Group / Task Force |
| Annika Auranen | Wellbeing Services County of Pirkanmaa, Tampere, Finland | Clinical Group / Task Force |
| Hannu Uusitalo | Wellbeing Services County of Pirkanmaa, Tampere, Finland | Clinical Group / Task Force |
| Teea Salmi | Wellbeing Services County of Pirkanmaa, Tampere, Finland | Clinical Group / Task Force |
| Venla Kurra | Wellbeing Services County of Pirkanmaa, Tampere, Finland | Clinical Group / Task Force |
| Laura Kotaniemi-Talonen | Wellbeing Services County of Pirkanmaa, Tampere, Finland | Clinical Group / Task Force |
| Argyro Bizaki-Vallaskangas | Wellbeing Services County of Pirkanmaa, Tampere, Finland | Clinical Group / Task Force |
| Juha Rinne | Wellbeing Services County of Southwest Finland, Turku, Finland | Clinical Group / Task Force |
| Roosa Kallionpää | Wellbeing Services County of Southwest Finland, Turku, Finland | Clinical Group / Task Force |
| Markku Voutilainen | Wellbeing Services County of Southwest Finland, Turku, Finland | Clinical Group / Task Force |
| Antti Palomäki | Wellbeing Services County of Southwest Finland, Turku, Finland | Clinical Group / Task Force |
| Laura Pirilä | Wellbeing Services County of Southwest Finland, Turku, Finland | Clinical Group / Task Force |
| Riitta Lahesmaa | Wellbeing Services County of Southwest Finland, Turku, Finland | Clinical Group / Task Force |
| Kaj Metsärinne | Wellbeing Services County of Southwest Finland, Turku, Finland | Clinical Group / Task Force |
| Jenni Aittokallio | Wellbeing Services County of Southwest Finland, Turku, Finland | Clinical Group / Task Force |
| Klaus Elenius | Wellbeing Services County of Southwest Finland, Turku, Finland | Clinical Group / Task Force |
| Sirkku Peltonen | Wellbeing Services County of Southwest Finland, Turku, Finland | Clinical Group / Task Force |
| Leena Koulu | Wellbeing Services County of Southwest Finland, Turku, Finland | Clinical Group / Task Force |
| Ulvi Gursoy | Wellbeing Services County of Southwest Finland, Turku, Finland | Clinical Group / Task Force |
| Varpu Jokimaa | Wellbeing Services County of Southwest Finland, Turku, Finland | Clinical Group / Task Force |
| Tytti Willberg | Wellbeing Services County of Southwest Finland, Turku, Finland | Clinical Group / Task Force |
| Adam Ziemann | Abbvie, Chicago, IL, United States | Clinical Group / Task Force |
| Nizar Smaoui | Abbvie, Chicago, IL, United States | Clinical Group / Task Force |
| Anne Lehtonen | Abbvie, Chicago, IL, United States | Clinical Group / Task Force |
| Apinya Lertratanakul | Abbvie, Chicago, IL, United States | Clinical Group / Task Force |
| Relja Popovic | Abbvie, Chicago, IL, United States | Clinical Group / Task Force |
| Mengzhen Liu | Abbvie, Chicago, IL, United States | Clinical Group / Task Force |
| Anneke Den Hollander | AbbVie, Chicago, IL, United States | Clinical Group / Task Force |
| Jan Freudenberg | AbbVie, Chicago, IL, United States | Clinical Group / Task Force |
| Britney Milkovich | AbbVie, Chicago, IL, United States | Clinical Group / Task Force |
| Andrew Blumenfeld | AbbVie, Chicago, IL, United States | Clinical Group / Task Force |
| Tushar Kumar | AbbVie, Chicago, IL, United States | Clinical Group / Task Force |
| Dirk Paul | AstraZeneca, Cambridge, United Kingdom | Clinical Group / Task Force |
| Bram Prins | AstraZeneca, Cambridge, United Kingdom | Clinical Group / Task Force |
| Eleanor Wheeler | AstraZeneca, Cambridge, United Kingdom | Clinical Group / Task Force |
| Kousik Kundu | AstraZeneca, Cambridge, United Kingdom | Clinical Group / Task Force |
| Santosh Atanur | AstraZeneca, Cambridge, United Kingdom | Clinical Group / Task Force |
| Andrew Lowe | AstraZeneca, Cambridge, United Kingdom | Clinical Group / Task Force |
| Thomas Spargo | AstraZeneca, Cambridge, United Kingdom | Clinical Group / Task Force |
| Oliver Burren | AstraZeneca, Cambridge, United Kingdom | Clinical Group / Task Force |
| Margarete Fabre | AstraZeneca, Cambridge, United Kingdom | Clinical Group / Task Force |
| Fabio Baschiera | Bayer AG, Leverkusen, Germany | Clinical Group / Task Force |
| Hans van Leeuwen | Bayer AG, Leverkusen, Germany | Clinical Group / Task Force |
| Himanshu Manchanda | Bayer AG, Leverkusen, Germany | Clinical Group / Task Force |
| Karl Heilbron | Bayer AG, Leverkusen, Germany | Clinical Group / Task Force |
| Martin Rao | Bayer AG, Leverkusen, Germany | Clinical Group / Task Force |
| Nicole Schmidt | Bayer AG, Leverkusen, Germany | Clinical Group / Task Force |
| Samu Kurki | Bayer AG, Leverkusen, Germany | Clinical Group / Task Force |
| Johanna Mielke | Bayer AG, Leverkusen, Germany | Clinical Group / Task Force |
| Juho Immonen | Bayer AG, Leverkusen, Germany | Clinical Group / Task Force |
| Thomas Battram | Bayer AG, Leverkusen, Germany | Clinical Group / Task Force |
| Tobias Hogrebe | Bayer AG, Leverkusen, Germany | Clinical Group / Task Force |
| Susan Eaton | Biogen, Cambridge, MA, United States | Clinical Group / Task Force |
| Ketian Yu | Biogen, Cambridge, MA, United States | Clinical Group / Task Force |
| Stephanie Loomis | Biogen, Cambridge, MA, United States | Clinical Group / Task Force |
| Coro Paisan-Ruiz | Biogen, Cambridge, MA, United States | Clinical Group / Task Force |
| Elke Markert | Boehringer Ingelheim, Ingelheim am Rhein, Germany | Clinical Group / Task Force |
| Frank Li | Boehringer Ingelheim, Ingelheim am Rhein, Germany | Clinical Group / Task Force |
| Yao Hu | Boehringer Ingelheim, Ingelheim am Rhein, Germany | Clinical Group / Task Force |
| Christoph Ogris | Boehringer Ingelheim, Ingelheim am Rhein, Germany | Clinical Group / Task Force |
| Eric Simon | Boehringer Ingelheim, Ingelheim am Rhein, Germany | Clinical Group / Task Force |
| Julio Cesar Bolivar Lopez | Boehringer Ingelheim, Ingelheim am Rhein, Germany | Clinical Group / Task Force |
| Monika Frysz | Boehringer Ingelheim, Ingelheim am Rhein, Germany | Clinical Group / Task Force |
| Marla Hochfeld | Bristol Myers Squibb, New York, NY, United States | Clinical Group / Task Force |
| Cara Carty | Bristol Myers Squibb, New York, NY, United States | Clinical Group / Task Force |
| Michael Turchin | Bristol Myers Squibb, New York, NY, United States | Clinical Group / Task Force |

| | | |
|---|---|---|
| Neelakshi Jog | Bristol Myers Squibb, New York, NY, United States | Clinical Group / Task Force |
| Corneliu Bodea | Bristol Myers Squibb, New York, NY, United States | Clinical Group / Task Force |
| Janie Shelton | Bristol Myers Squibb, New York, NY, United States | Clinical Group / Task Force |
| Chen Li | Bristol Myers Squibb, New York, NY, United States | Clinical Group / Task Force |
| Kritika Singh | Bristol Myers Squibb, New York, NY, United States | Clinical Group / Task Force |
| Peng Jiang | Bristol Myers Squibb, New York, NY, United States | Clinical Group / Task Force |
| Stephanie Loomis | Bristol Myers Squibb, New York, NY, United States | Clinical Group / Task Force |
| Elena Sanchez | Bristol Myers Squibb, New York, NY, United States | Clinical Group / Task Force |
| Lilith Moss | Bristol Myers Squibb, New York, NY, United States | Clinical Group / Task Force |
| Zijie Zhao | Bristol Myers Squibb, New York, NY, United States | Clinical Group / Task Force |
| Anna Podgornaia | Bristol Myers Squibb, New York, NY, United States | Clinical Group / Task Force |
| Natalie Bowers | Genentech, San Francisco, CA, United States | Clinical Group / Task Force |
| Edmond Teng | Genentech, San Francisco, CA, United States | Clinical Group / Task Force |
| Tim Lu | Genentech, San Francisco, CA, United States | Clinical Group / Task Force |
| Hubert Chen | Genentech, San Francisco, CA, United States | Clinical Group / Task Force |
| Jennifer Schutzman | Genentech, San Francisco, CA, United States | Clinical Group / Task Force |
| Erich Strauss | Genentech, San Francisco, CA, United States | Clinical Group / Task Force |
| Hao Chen | Genentech, San Francisco, CA, United States | Clinical Group / Task Force |
| David Choy | Genentech, San Francisco, CA, United States | Clinical Group / Task Force |
| Rion Pendergrass | Genentech, San Francisco, CA, United States | Clinical Group / Task Force |
| Brian Yaspan | Genentech, San Francisco, CA, United States | Clinical Group / Task Force |
| Cameron Adams | Genentech, San Francisco, CA, United States | Clinical Group / Task Force |
| Mark McCarthy | Genentech, San Francisco, CA, United States | Clinical Group / Task Force |
| Michael Rothenberg | Genentech, San Francisco, CA, United States | Clinical Group / Task Force |
| Rion Pendergrass | Genentech, San Francisco, CA, United States | Clinical Group / Task Force |
| Sergio Dellepiane | Genentech, San Francisco, CA, United States | Clinical Group / Task Force |
| Anubha Mahajan | Genentech, San Francisco, CA, United States | Clinical Group / Task Force |
| Michael Holmes | Genentech, San Francisco, CA, United States | Clinical Group / Task Force |
| Anubha Mahajan | Genentech, San Francisco, CA, United States | Clinical Group / Task Force |
| Diana Chang | Genentech, San Francisco, CA, United States | Clinical Group / Task Force |
| Tushar Bhangale | Genentech, San Francisco, CA, United States | Clinical Group / Task Force |
| Fanli Xu | GlaxoSmithKline, Brentford, United Kingdom | Clinical Group / Task Force |
| Laura Addis | GlaxoSmithKline, Brentford, United Kingdom | Clinical Group / Task Force |
| John Eicher | GlaxoSmithKline, Brentford, United Kingdom | Clinical Group / Task Force |
| Linda McCarthy | GlaxoSmithKline, Brentford, United Kingdom | Clinical Group / Task Force |
| Jorge Esparza Gordillo | GlaxoSmithKline, Brentford, United Kingdom | Clinical Group / Task Force |
| Joanna Betts | GlaxoSmithKline, Brentford, United Kingdom | Clinical Group / Task Force |
| Rajashree Mishra | GlaxoSmithKline, Brentford, United Kingdom | Clinical Group / Task Force |
| Audrey Chu | GlaxoSmithKline, Brentford, United Kingdom | Clinical Group / Task Force |
| Diptee Kulkarni | GlaxoSmithKline, Brentford, United Kingdom | Clinical Group / Task Force |
| Janet Kumar | GlaxoSmithKline, Collegeville, PA, United States | Clinical Group / Task Force |
| Charli Harlow | GlaxoSmithKline, Collegeville, PA, United States | Clinical Group / Task Force |
| Lea Sarow-Blat | GlaxoSmithKline, Collegeville, PA, United States | Clinical Group / Task Force |
| Diana L.Cousminer | GlaxoSmithKline, Collegeville, PA, United States | Clinical Group / Task Force |
| Jagtar Nijjar | GlaxoSmithKline, Collegeville, PA, United States | Clinical Group / Task Force |
| Jessica Chao | GlaxoSmithKline, Collegeville, PA, United States | Clinical Group / Task Force |
| Michal Magid | GlaxoSmithKline, Collegeville, PA, United States | Clinical Group / Task Force |
| Shashank Jariwala | GlaxoSmithKline, Collegeville, PA, United States | Clinical Group / Task Force |
| Chris Floyd | GlaxoSmithKline, Collegeville, PA, United States | Clinical Group / Task Force |
| Dan Swerdlow | GlaxoSmithKline, Collegeville, PA, United States | Clinical Group / Task Force |
| Erding Hu | GlaxoSmithKline, Collegeville, PA, United States | Clinical Group / Task Force |
| Prerak Desai | GlaxoSmithKline, Collegeville, PA, United States | Clinical Group / Task Force |
| Stephen Haddad | GlaxoSmithKline, Collegeville, PA, United States | Clinical Group / Task Force |
| Damien Croteau-Chonka | GlaxoSmithKline, Collegeville, PA, United States | Clinical Group / Task Force |
| Billy Fahy | GlaxoSmithKline, Collegeville, PA, United States | Clinical Group / Task Force |
| Paola Bronson | GlaxoSmithKline, Collegeville, PA, United States | Clinical Group / Task Force |
| Kirsi Auro | GlaxoSmithKline, Espoo, Finland | Clinical Group / Task Force |
| David Pulford | GlaxoSmithKline, Stevenage, United Kingdom | Clinical Group / Task Force |
| Sauli Vuoti | Janssen-Cilag Oy, Espoo, Finland | Clinical Group / Task Force |
| Dermot Reilly | Johnson & Johnson Innovative Medicine, Boston, MA, United States | Clinical Group / Task Force |
| Karen He | Johnson & Johnson Innovative Medicine, Spring House, PA, United States | Clinical Group / Task Force |
| Ekaterina Khramtsova | Johnson & Johnson Innovative Medicine, Spring House, PA, United States | Clinical Group / Task Force |
| Amy Hart | Johnson & Johnson Innovative Medicine, Spring House, PA, United States | Clinical Group / Task Force |
| Meijian Guan | Johnson & Johnson Innovative Medicine, Spring House, PA, United States | Clinical Group / Task Force |
| Alessandro Porello | Johnson & Johnson Innovative Medicine, Spring House, PA, United States | Clinical Group / Task Force |
| P. Dunnmon | Johnson & Johnson Innovative Medicine, Spring House, PA, United States | Clinical Group / Task Force |
| Sara Gale | Johnson & Johnson Innovative Medicine, Spring House, PA, United States | Clinical Group / Task Force |
| Brice Keyes | Johnson & Johnson Innovative Medicine, Spring House, PA, United States | Clinical Group / Task Force |
| John Kwon | Johnson & Johnson Innovative Medicine, Spring House, PA, United States | Clinical Group / Task Force |
| Jonathan Sherlock | Johnson & Johnson Innovative Medicine, Spring House, PA, United States | Clinical Group / Task Force |
| Matt Loza | Johnson & Johnson Innovative Medicine, Spring House, PA, United States | Clinical Group / Task Force |
| Chris Whelan | Johnson & Johnson Innovative Medicine, Spring House, PA, United States | Clinical Group / Task Force |
| W Galpern | Johnson & Johnson Innovative Medicine, Spring House, PA, United States | Clinical Group / Task Force |
| Yanfei Zhang | Johnson & Johnson Innovative Medicine, Spring House, PA, United States | Clinical Group / Task Force |
| Mona Selej | Johnson & Johnson Innovative Medicine, Spring House, PA, United States | Clinical Group / Task Force |
| Abolfazl Doostparast Torshizi | Johnson & Johnson Innovative Medicine, Spring House, PA, United States | Clinical Group / Task Force |
| Qingqin S Li | Johnson & Johnson Innovative Medicine, Titusville, NJ, United States | Clinical Group / Task Force |
| Sahar Mozzafari | Maze Therapeutics, San Francisco, CA, United States | Clinical Group / Task Force |
| Christopher Deboever | Maze Therapeutics, San Francisco, CA, United States | Clinical Group / Task Force |
| Jason Miller | Merck, Kenilworth, NJ, United States | Clinical Group / Task Force |
| Fabiana Farias | Merck, Kenilworth, NJ, United States | Clinical Group / Task Force |
| Andrey Loboda | Merck, Kenilworth, NJ, United States | Clinical Group / Task Force |
| Jorge Del-aguila | Merck, Kenilworth, NJ, United States | Clinical Group / Task Force |
| Elisabeth Vollmann | Merck, Kenilworth, NJ, United States | Clinical Group / Task Force |
| Jozsef Karman | Merck, Kenilworth, NJ, United States | Clinical Group / Task Force |
| Julie Fiore | Merck, Kenilworth, NJ, United States | Clinical Group / Task Force |
| Rajesh Kamath | Merck, Kenilworth, NJ, United States | Clinical Group / Task Force |
| Andrei Popescu | Merck, Kenilworth, NJ, United States | Clinical Group / Task Force |
| Delphine Fagegaltier | Merck, Kenilworth, NJ, United States | Clinical Group / Task Force |
| Travis Barr | Merck, Kenilworth, NJ, United States | Clinical Group / Task Force |
| Aristide Merola | Merck, Kenilworth, NJ, United States | Clinical Group / Task Force |
| Oliver Freeman | Merck, Kenilworth, NJ, United States | Clinical Group / Task Force |
| Simonne Longerich | Merck, Kenilworth, NJ, United States | Clinical Group / Task Force |
| Enrico Ferrero | Novartis Institutes for BioMedical Research, Cambridge, MA, United States | Clinical Group / Task Force |
| Nikos Patsopoulos | Novartis Institutes for BioMedical Research, Cambridge, MA, United States | Clinical Group / Task Force |
| Nancy Finkel | Novartis Institutes for BioMedical Research, Cambridge, MA, United States | Clinical Group / Task Force |
| Sabina Pfister | Novartis Institutes for BioMedical Research, Cambridge, MA, United States | Clinical Group / Task Force |
| Shola Richards | Novartis Institutes for BioMedical Research, Cambridge, MA, United States | Clinical Group / Task Force |
| Katherine Mccauley | Novartis Institutes for BioMedical Research, Cambridge, MA, United States | Clinical Group / Task Force |
| Xiaobo Xia | Novartis Institutes for BioMedical Research, Cambridge, MA, United States | Clinical Group / Task Force |
| Mike Mendelson | Novartis Institutes for BioMedical Research, Cambridge, MA, United States | Clinical Group / Task Force |
| Majd Mouded | Novartis, Basel, Switzerland | Clinical Group / Task Force |
| Debby Ngo | Novartis, Basel, Switzerland | Clinical Group / Task Force |
| Kirsi Kalpala | Pfizer, New York, NY, United States | Clinical Group / Task Force |
| Melissa Miller | Pfizer, New York, NY, United States | Clinical Group / Task Force |
| Nan Bing | Pfizer, New York, NY, United States | Clinical Group / Task Force |
| Jaakko Parkkinen | Pfizer, New York, NY, United States | Clinical Group / Task Force |

| | | | |
|---|---|---|---|
| Heli Lehtonen | Pfizer, New York, NY, United States | **Clinical Group / Task Force** | |
| Stefan McDonough | Pfizer, New York, NY, United States | **Clinical Group / Task Force** | |
| Ying Wu | Pfizer, New York, NY, United States | **Clinical Group / Task Force** | |
| Erin Macdonald-Dunlop | Pfizer, New York, NY, United States | **Clinical Group / Task Force** | |
| Jessica Chung | Pfizer, New York, NY, United States | **Clinical Group / Task Force** | |
| Michael McLean | Pfizer, New York, NY, United States | **Clinical Group / Task Force** | |
| Joshua Chiou | Pfizer, New York, NY, United States | **Clinical Group / Task Force** | |
| Hye In Kim | Pfizer, New York, NY, United States | **Clinical Group / Task Force** | |
| Sivakumar Pitchumani | Pfizer, New York, NY, United States | **Clinical Group / Task Force** | |
| Sumedha Jassal | Pfizer, New York, NY, United States | **Clinical Group / Task Force** | |
| Madhurima Saxena | Pfizer, New York, NY, United States | **Clinical Group / Task Force** | |
| Catherine O'Riordan | Translational Sciences, Sanofi R&D, Framingham, MA, USA | **Clinical Group / Task Force** | |
| Samuel Lessard | Translational Sciences, Sanofi R&D, Framingham, MA, USA | **Clinical Group / Task Force** | |
| Suzanne Jacobs | Translational Sciences, Sanofi R&D, Framingham, MA, USA | **Clinical Group / Task Force** | |
| Hamid Mattoo | Translational Sciences, Sanofi R&D, Framingham, MA, USA | **Clinical Group / Task Force** | |
| David Habiel | Translational Sciences, Sanofi R&D, Framingham, MA, USA | **Clinical Group / Task Force** | |
| Guanling Huan | Translational Sciences, Sanofi R&D, Framingham, MA, USA | **Clinical Group / Task Force** | |
| Lila Kallio | Auria Biobank / University of Turku / Wellbeing Services County of Southwest Finland, Turku, Finland | **Biobank directors** | |
| Tiina Wahlfors | THL Biobank / Finnish Institute for Health and Welfare (THL), Helsinki, Finland | **Biobank directors** | |
| Jukka Partanen | Finnish Red Cross Blood Service / Finnish Hematology Registry and Clinical Biobank, Helsinki, Finland | **Biobank directors** | |
| Eero Punkka | Helsinki Biobank / Helsinki University and Hospital District of Helsinki and Uusimaa, Helsinki | **Biobank directors** | |
| Raisa Serpi | Northern Finland Biobank Borealis / University of Oulu / Wellbeing services county of North Ostrobothnia, Oulu, Finland | **Biobank directors** | |
| Sanna Siltanen | Finnish Clinical Biobank Tampere / University of Tampere / Wellbeing Services County of Pirkanmaa, Tampere, Finland | **Biobank directors** | |
| Veli-Matti Kosma | Biobank of Eastern Finland / University of Eastern Finland / Wellbeing services county of North Savo, Kuopio, Finland | **Biobank directors** | |
| Tiina Jokela | Central Finland Biobank / University of Jyväskylä / Wellbeing Services County of Central Finland, Jyväskylä, Finland | **Biobank directors** | |
| Anu Jalanko | Institute for Molecular Medicine Finland (FIMM), HiLIFE, University of Helsinki, Helsinki, Finland | **FinnGen Teams** | **Administration** |
| Risto Kajanne | Institute for Molecular Medicine Finland (FIMM), HiLIFE, University of Helsinki, Helsinki, Finland | **FinnGen Teams** | **Administration** |
| Mervi Aavikko | Institute for Molecular Medicine Finland (FIMM), HiLIFE, University of Helsinki, Helsinki, Finland | **FinnGen Teams** | **Administration** |
| Helen Cooper | Institute for Molecular Medicine Finland (FIMM), HiLIFE, University of Helsinki, Helsinki, Finland | **FinnGen Teams** | **Administration** |
| Denise Öller | Institute for Molecular Medicine Finland (FIMM), HiLIFE, University of Helsinki, Helsinki, Finland | **FinnGen Teams** | **Administration** |
| Tarja Laitinen | Institute for Molecular Medicine Finland (FIMM), HiLIFE, University of Helsinki, Helsinki, Finland | **FinnGen Teams** | **Administration** |
| Rodos Rodosthenous | Institute for Molecular Medicine Finland (FIMM), HiLIFE, University of Helsinki, Helsinki, Finland | **FinnGen Teams** | **Administration** |
| Sofia Kuitunen | University of Helsinki, Helsinki, Finland | **FinnGen Teams** | **Administration** |
| Mitja Kurki | Institute for Molecular Medicine Finland (FIMM), HiLIFE, University of Helsinki, Helsinki, Finland; Broad Institute, Cambridge, MA, United States | **FinnGen Teams** | **Analysis** |
| Juha Karjalainen | Institute for Molecular Medicine Finland (FIMM), HiLIFE, University of Helsinki, Helsinki, Finland | **FinnGen Teams** | **Analysis** |
| Pietro Della Briotta Parolo | Institute for Molecular Medicine Finland (FIMM), HiLIFE, University of Helsinki, Helsinki, Finland | **FinnGen Teams** | **Analysis** |
| Arto Lehisto | Institute for Molecular Medicine Finland (FIMM), HiLIFE, University of Helsinki, Helsinki, Finland | **FinnGen Teams** | **Analysis** |
| Juha Mehtonen | Institute for Molecular Medicine Finland (FIMM), HiLIFE, University of Helsinki, Helsinki, Finland | **FinnGen Teams** | **Analysis** |
| Reza Jabal | Institute for Molecular Medicine Finland (FIMM), HiLIFE, University of Helsinki, Helsinki, Finland; Broad Institute, Cambridge, MA, United States | **FinnGen Teams** | **Analysis** |
| Mutaamba Maasha | Institute for Molecular Medicine Finland (FIMM), HiLIFE, University of Helsinki, Helsinki, Finland; Broad Institute, Cambridge, MA, United States | **FinnGen Teams** | **Analysis** |
| Sanni Ruotsalainen | Institute for Molecular Medicine Finland (FIMM), HiLIFE, University of Helsinki, Helsinki, Finland | **FinnGen Teams** | **Analysis** |
| Samuel Jones | Institute for Molecular Medicine Finland (FIMM), HiLIFE, University of Helsinki, Helsinki, Finland | **FinnGen Teams** | **Analysis** |
| Raymond Walters | Institute for Molecular Medicine Finland (FIMM), HiLIFE, University of Helsinki, Helsinki, Finland; Broad Institute, Cambridge, MA, United States | **FinnGen Teams** | **Analysis** |
| Paavo Häppölä | Institute for Molecular Medicine Finland (FIMM), HiLIFE, University of Helsinki, Helsinki, Finland | **FinnGen Teams** | **Analysis** |
| Topi Paavilainen | Institute for Molecular Medicine Finland (FIMM), HiLIFE, University of Helsinki, Helsinki, Finland | **FinnGen Teams** | **Analysis** |
| L. Elisa Lahtela | Institute for Molecular Medicine Finland (FIMM), HiLIFE, University of Helsinki, Helsinki, Finland | **FinnGen Teams** | **Disease Task Forces** |
| Johanna Paltta | Institute for Molecular Medicine Finland (FIMM), HiLIFE, University of Helsinki, Helsinki, Finland; University of Turku, Turku, Finland | **FinnGen Teams** | **Disease Task Forces** |
| Juulia Partanen | Institute for Molecular Medicine Finland (FIMM), HiLIFE, University of Helsinki, Helsinki, Finland | **FinnGen Teams** | **Disease Task Forces** |
| Olli K. Pietiläinen | Institute for Molecular Medicine Finland (FIMM), HiLIFE, University of Helsinki, Helsinki, Finland | **FinnGen Teams** | **Disease Task Forces** |
| Veera Timonen | Institute for Molecular Medicine Finland (FIMM), HiLIFE, University of Helsinki, Helsinki, Finland | **FinnGen Teams** | **Disease Task Forces** |
| Mari Kaunisto | Institute for Molecular Medicine Finland (FIMM), HiLIFE, University of Helsinki, Helsinki, Finland | **FinnGen Teams** | **Communication** |
| Elina Kilpeläinen | Institute for Molecular Medicine Finland (FIMM), HiLIFE, University of Helsinki, Helsinki, Finland | **FinnGen Teams** | **Sandbox & Cloud Services** |
| Tianduanyi Wang | Institute for Molecular Medicine Finland (FIMM), HiLIFE, University of Helsinki, Helsinki, Finland | **FinnGen Teams** | **Sandbox & Cloud Services** |
| Timo P. Sipilä | Institute for Molecular Medicine Finland (FIMM), HiLIFE, University of Helsinki, Helsinki, Finland | **FinnGen Teams** | **Sandbox & Cloud Services** |
| Oluwaseun Alexander Dada | Institute for Molecular Medicine Finland (FIMM), HiLIFE, University of Helsinki, Helsinki, Finland | **FinnGen Teams** | **Sandbox & Cloud Services** |
| Awaisa Ghazal | Institute for Molecular Medicine Finland (FIMM), HiLIFE, University of Helsinki, Helsinki, Finland | **FinnGen Teams** | **Sandbox & Cloud Services** |
| Rigbe Weldatsadik | Institute for Molecular Medicine Finland (FIMM), HiLIFE, University of Helsinki, Helsinki, Finland | **FinnGen Teams** | **Sandbox & Cloud Services** |
| Jaska Uimonen | Institute for Molecular Medicine Finland (FIMM), HiLIFE, University of Helsinki, Helsinki, Finland | **FinnGen Teams** | **Sandbox & Cloud Services** |
| Kati Donner | Institute for Molecular Medicine Finland (FIMM), HiLIFE, University of Helsinki, Helsinki, Finland | **FinnGen Teams** | **Genotyping** |
| Anu Loukola | Helsinki Biobank / Helsinki University and Hospital District of Helsinki and Uusimaa, Helsinki | **FinnGen Teams** | **Sample Collection Coordination** |
| Päivi Laiho | THL Biobank / Finnish Institute for Health and Welfare (THL), Helsinki, Finland | **FinnGen Teams** | **Sample Logistics** |
| Susanna Lemmelä | Institute for Molecular Medicine Finland (FIMM), HiLIFE, University of Helsinki, Helsinki, Finland | **FinnGen Teams** | **Register data and sample logistics** |
| Teemu Paajanen | THL Biobank / Finnish Institute for Health and Welfare (THL), Helsinki, Finland | **FinnGen Teams** | **Register data and sample logistics** |
| Arto Pietilä | THL Biobank / Finnish Institute for Health and Welfare (THL), Helsinki, Finland | **FinnGen Teams** | **Register data and sample logistics** |
| Aki Havulinna | THL Biobank / Finnish Institute for Health and Welfare (THL), Helsinki, Finland | **FinnGen Teams** | **Register data and sample logistics** |

| | | | |
|---|---|---|---|
| Auli Toivola | Institute for Molecular Medicine Finland (FIMM), HiLIFE, University of Helsinki, Helsinki, Finland | **FinnGen Teams** | **Register data and sample logistics** |
| Kristina Zguro | Institute for Molecular Medicine Finland (FIMM), HiLIFE, University of Helsinki, Helsinki, Finland | **FinnGen Teams** | **Register data and sample logistics** |
| Mary Pat Reeve | Institute for Molecular Medicine Finland (FIMM), HiLIFE, University of Helsinki, Helsinki, Finland; Broad Institute, Cambridge, MA, United States | **FinnGen Teams** | **Phenotype team** |
| Shanmukha Sampath Padmanabh | Institute for Molecular Medicine Finland (FIMM), HiLIFE, University of Helsinki, Helsinki, Finland | **FinnGen Teams** | **Phenotype team** |
| Harri Siirtola | University of Tampere, Tampere, Finland | **FinnGen Teams** | **Phenotype team** |
| Javier Gracia-Tabuenca | University of Tampere, Tampere, Finland | **FinnGen Teams** | **Phenotype team** |
| Marika Kaakinen | Institute for Molecular Medicine Finland (FIMM), HiLIFE, University of Helsinki, Helsinki, Finland | **FinnGen Teams** | **Phenotype team** |
| Shuang Luo | Institute for Molecular Medicine Finland (FIMM), HiLIFE, University of Helsinki, Helsinki, Finland | **FinnGen Teams** | **Phenotype team** |
| Vincent Llorens | Institute for Molecular Medicine Finland (FIMM), HiLIFE, University of Helsinki, Helsinki, Finland | **FinnGen Teams** | **Phenotype team** |
| Dawit Yohannes | Institute for Molecular Medicine Finland (FIMM), HiLIFE, University of Helsinki, Helsinki, Finland | **FinnGen Teams** | **Phenotype team** |
| Iina Laak | Institute for Molecular Medicine Finland (FIMM), HiLIFE, University of Helsinki, Helsinki, Finland | **FinnGen Teams** | **Data protection officer** |
| Mervi Ahlroth | Finnish Biobank Cooperative - FINBB | **FinnGen Teams** | **FINBB - Finnish biobank cooperative** |
| Johanna Mäkelä | Finnish Biobank Cooperative - FINBB | **FinnGen Teams** | **FINBB - Finnish biobank cooperative** |
| Pauli Wihuri | Finnish Biobank Cooperative - FINBB | **FinnGen Teams** | **FINBB - Finnish biobank cooperative** |
| Tom Southerington | Finnish Biobank Cooperative - FINBB | **FinnGen Teams** | **FINBB - Finnish biobank cooperative** |
| Meri Lähteenmäki | Finnish Biobank Cooperative - FINBB | **FinnGen Teams** | **FINBB - Finnish biobank cooperative** |
